# The emergence and persistence of inequalities in adolescent mental health: the Resilience, Ethnicity and AdolesCent Mental Health (REACH) cohorts

**DOI:** 10.1101/2025.01.07.25320111

**Authors:** Gemma Knowles, Charlotte Gayer-Anderson, Samantha Davis, Daniel Stanyon, Aisha Ofori, Rachel Blakey, Thai-sha Richards, Adna Hashi, Karima Shyan Clement-Gbede, Jonas Kitisu, Niiokani Tettey, Katie Lowis, Alice Turner, Lynsey Dorn, Esther Putzgruber, Vanessa Pinfold, Seeromanie Harding, Kamaldeep Bhui, Craig Morgan

## Abstract

**Background:** Mental health problems are not distributed equally in society. Our understanding of when social inequalities in mental health emerge is limited. We sought to examine inequalities in trajectories of mental distress in diverse, representative cohorts of adolescents in inner-London.

**Methods:** We analysed longitudinal data from our cohort study of adolescent mental health, REACH (n=4663; 51% girls, 29% free school meals [FSM], 85% minoritised ethnic groups). We used latent growth curve models to estimate trajectories of mental distress (total, internalising, and externalising scores from the self-report Strengths and Difficulties Questionnaire) from age 11-16 years, overall and by gender, FSM, ethnic group, and their intersections.

**Results:** We found strong evidence of differences in trajectories of mental distress by gender and FSM. Higher mean internalising scores in girls (vs. boys) were evident at age 11-12 and this inequality widened year-on-year (difference in mean intercepts: 0.74 [95% CI 0.52, 0.96]; slopes: 0.50 [0.39, 0.61]). Higher mean levels of distress among those receiving FSM (vs. not) were evident at age 11-12 years (e.g., difference in intercepts, general distress: 0.79 [0.19, 1.39]), and this difference, though modest, persisted through adolescence. By ethnic group and intersecting identities, the picture was more complex and mixed. Broadly, Black African youth generally reported better mental health trajectories vs. their peers; Black Caribbean and Mixed Black-and-White youth shared similar trajectories, differing somewhat from Black African; and by age 16, internalising distress was highest among lower-income White British girls.

**Conclusions:** In diverse inner-cities, adolescence is an important period in the emergence and persistence of some of the inequalities in mental health reported in adults; others are more nuanced.

## INTRODUCTION

Among adults in the UK, there is robust evidence of marked inequalities in mental health by social group. For instance, depression and anxiety are more common among women compared with men (McManus et al., 2016); substance use disorders and deaths by suicide tend to be more common among men compared with women (Office for National Statistics, 2022; McManus et al., 2016); there are clear socioeconomic gradients in almost all types of mental disorder (Allen et al., 2014); and there are marked inequalities in (some types of) mental health problems – and in the use of involuntary psychiatric detentions – by ethnic group (Bhui et al., 2003; Morgan et al., 2006).

However, our understanding of when, during development, inequalities in mental distress first emerge, what causes them, and how and when to intervene to prevent and mitigate them, is limited. In part this is because the available UK-based data on child and adolescent mental health are limited in notable ways. First, most existing evidence is based on nationally representative and/or predominantly White samples (Sadler et al., 2018; Patalay & Gage, 2019) and there is a distinct lack of data on representative samples of young people from diverse backgrounds (Knowles et al., 2021). Second, much of the evidence is cross-sectional (Sadler et al., 2018) or is longitudinal but with too few (Clark et al., 2007; Maynard et al., 2007) or too infrequent (McElroy et al., 2023) timepoints to understand in detail the onset and course of inequalities in mental distress during periods of rapid developmental change, e.g., adolescence, the period in which many mental health problems first emerge (Jones, 2013; Kessler et al., 2007). Third, where data on diverse groups of young people do exist it is generally outdated, e.g., relates to the experience of those born in the late 1980s and early 1990s (Clark et al., 2007; Karamanos et al., 2021). A lot has changed since then, and there is evidence of rising rates of and growing inequalities in mental distress in more recent compared with prior generations of young people (Collishaw et al., 2019; McElroy et al., 2023). Fourth, most research has relied fully or primarily on parent-report data on young people’s mental health (Terhaag et al., 2021; Sadler et al., 2018, McElroy et al., 2023), which may be less accurate in adolescence as young people develop independence (Booth et al., 2023).

These caveats noted, there is somewhat consistent evidence in some areas. For example: (a) strong evidence of higher prevalence of internalising/emotional problems in girls compared with boys from around age 14 (Terhaag et al., 2021; Sadler et al., 2018; McElroy et al., 2023), but mixed evidence on gender inequalities in externalising problems (Murray et al., 2022; Yoon et al., 2022; Blakey et al., 2021; McElroy et al., 2023; Sadler et al., 2018), (b) strong evidence of the negative impacts of poverty on mental health in childhood (Wickham et al., 2017), and (c) some evidence of a ‘mental health advantage’ among young people from ‘minority ethnic groups’ – particularly Indian and Black African – compared with their white British peers (Goodman et al., 2008; Sadler et al., 2018). On the latter, however, the evidence is mixed (e.g., some point to similar levels of distress in Black Caribbean and White youth in mid-adolescence (Bains & Gutman, 2021)) and tends to be limited by unrepresentative samples and broad groupings (e.g., White vs ‘minority’) which conceal vast differences in culture, experience, and social and historic context within groups in the UK.

Importantly, we also know very little about the mental health trajectories of young people with multiple marginalised identities (Kern et al., 2020; Maynard et al., 2007; Patil et al., 2018). Intersectionality theory provides a lens for understanding the impacts of overlapping and interlocking forms of discrimination on the lives of groups and individuals with multiple marginalised – and multiple privileged – identities (Crenshaw, 1989). While a truly intersectional approach remains methodologically challenging in developmental and lifecourse epidemiology (Bauer et al., 2021), the importance of at least considering lifecourse trajectories beyond single-identity groupings is clear (Ghavami et al., 2016). For instance, in the US there is evidence that the education-related health benefits enjoyed by the ‘White majority’ are not shared by Black communities, in part because higher education does not return the same financial gains for Black communities (Shuey & Willson, 2008). Likewise, in the international literature, there is growing evidence that gender inequalities in adolescent mental health – previously considered to be broadly similar across populations – may vary in size, direction, and age of onset across time and place (Knowles at al., 2024; Campbell et al., 2021; Drosopoulou et al., 2023; Platt et al., 2021). Yet, our understanding of whether gender inequalities in mental health emerge and evolve differently across diverse social groups (e.g., income, ethnic group) within the UK is limited.

In this paper, we use longitudinal data from our accelerated cohort study of adolescent mental health, REACH (Resilience, Ethnicity, and AdolesCent Mental Health; Knowles et al., 2021), to examine the extent, nature, and course of social inequalities in trajectories of mental distress (specifically internalising and externalising problems) in young people from diverse backgrounds. The REACH cohorts are large, representative, and highly diverse, and those taking part are providing information annually through adolescence. As such, REACH addresses many of the limitations of the existing evidence on the development of social inequalities in mental health.

### Hypotheses

As discussed, there is limited and often mixed evidence on mental health trajectories in diverse groups of adolescents in the UK. Therefore, we set out to test four broad hypotheses based on the more consistent evidence (outlined above), our understanding of inequalities in mental health among UK-based adults (McManus et al., 2016), findings from analyses of REACH baseline data (Knowles et al., 2021), and input from our young co-researchers (see below and author-line):

1. Gender inequalities in internalising problems emerge in early adolescence and widen-year-on-year.
2. Young people from low-income households already experience worse mental health than their peers by the start of adolescence, and these inequalities persist with age.
3. Internalising scores are broadly similar across ethnic groups in early adolescence but diverge with age for some groups, e.g., lower in Black African compared with other groups.
4. Broadly, trajectories of distress are generally worst among those with multiple marginalised identities.

## METHODS

### Study design

REACH is an accelerated cohort study of adolescent mental health in inner-London. Three cohorts of young people are taking part: Cohort 1, age 11-12 years (school year 7) at baseline; Cohort 2, age 12-13 years (school year 8) at baseline; and Cohort 3, age 13-14 years (school year 9) at baseline. The study design is illustrated in Figure S1 and in the Cohort Profile (Knowles et al., 2022). This paper uses data from the first three waves of REACH, collected between January 2016 and March 2020.

### Setting, participants

Participants were recruited from twelve state-funded secondary schools in Lambeth and Southwark. These boroughs are among the most densely populated and diverse areas in England, and prevalence of mental health problems in adults is higher here than reported nationally (Hatch et al., 2012). Schools were recruited using a quota sampling approach (see Knowles et al., 2022) to generate cohorts that are highly representative of the target population based on demographics, i.e., age, sex/gender, ethnic group, and FSM. At the twelve participating schools, all students in school years 7 to 9 (n, 4945) were invited to take part at baseline.

### Procedures

Informed consent was obtained for all participants at each wave. Around two weeks prior to data collection, researchers delivered in-school talks about REACH to students and staff and provided written information for students and their parents/carers. Parents were asked to return a form or contact the school or research team if they did not want their child to take part. On the day of data collection, students again received verbal and written information from researchers and provided written assent before completing computerised questionnaires. Trained researchers were present to support students and answer questions.

## Measures

### Mental health

At each wave and in each cohort, mental health was assessed with the widely used self-report Strengths and Difficulties Questionnaire (SDQ) for 11-17-year-olds (Goodman, 2001). The SDQ comprises 25 items spanning five subdomains: emotional problems, peer problems, hyperactivity, conduct problems, and prosocial behaviour. Each item has three response options: not true, somewhat true, certainly true (scored 0 to 2). Within each subdomain, the five items are summed to produce a score ranging from 0 to 10. The emotional and peer scores are summed to produce an ‘internalising score’ and conduct and hyperactivity an ‘externalising score’, each ranging from 0 to 20. A ‘total difficulties’ (general distress) score ranging from 0 to 40 is calculated by summing the emotional, peer, conduct, and hyperactivity scores. The higher-order internalising, externalising, and general distress scores used in this analysis are supported in community-based samples (Goodman et al., 2010). Higher scores indicate higher levels of distress.

### Demographic characteristics

Sex/gender, age, ethnic group, and FSM were self-reported. Participants were asked to select their ethnic group from the 18 categories used in the 2011 Census (Office for National Statistics, 2012). Five ethnic groups – the largest in the cohorts – were included in the analysis: Black African, Black Caribbean, Mixed Black-and-White, White British, and Other. The latter comprised a wide range of ethnic groups (e.g., Indian, Chinese, Arab) that, within each cohort, were too small to disaggregate. The ‘Other’ ethnic group, then, is highly heterogenous and not included in specific hypotheses.

### Data analysis

First, we examined the extent and predictors of missingness on all variables and calculated summary statistics, at each wave, overall and by cohort, gender, FSM, and ethnic group. Second, we examined the distributions and correlation/covariance matrix of the repeated measures of mental distress. Third, we performed a series of analyses to test the assumptions of the accelerated cohort design (see Supporting Information Appendix S1). Fourth, we used a series of latent growth curve models (LCMs) to estimate mean trajectories of distress overall and by group (path diagram in Figure 1). For each distress metric separately, we fit intercept-only (no-growth), linear- and quadratic-growth models, with heteroscedastic and homoscedastic residuals, and used standard model fit indices to determine the best fitting model. We took a holistic approach to evaluating model fit. That is, we considered all fit indices collectively, used broadly supported thresholds as a guide (i.e., TLI>0.95, CFI>0.95, RMSEA<0.06, SRMR<0.08 to indicate good fit), and gave less weight to indices that are sensitive to sample size (Hu & Bentler, 1999). We used the CLUSTER option (*Mplus*) and robust maximum likelihood estimation to account for clustering of participants within schools. In sensitivity analyses, we re-ran models with school as a fixed effect, given the small number of schools in the study, but this made no difference to the results (Table S3). We then added (a) gender (0=boy, 1=girl), (b) FSM (0=no, 1=yes), and (c) ethnic group as time invariant covariates in separate models and examined the strength of evidence for differences in mean intercepts and mean slopes by group. We then added interaction terms for gender*FSM, gender*ethnic group, FSM*ethnic group, and FSM*ethnic group within gender. *Missing data:* The extent of missing data on demographic characteristics was very low. Sex/gender was fully observed. FSM was missing for 65 participants (1.4%). Ethnic group was ‘unknown’ for seven participants (0.2%); we merged these into the ‘Other’ ethnic group. Full information maximum likelihood estimation was used to account for missing data in the repeated measures of mental health, under the missing at random (MAR) assumption. Analyses were conducted in Stata version 17 and Mplus version 8 (Muthén & Muthén, 2017).

**Figure 1.**
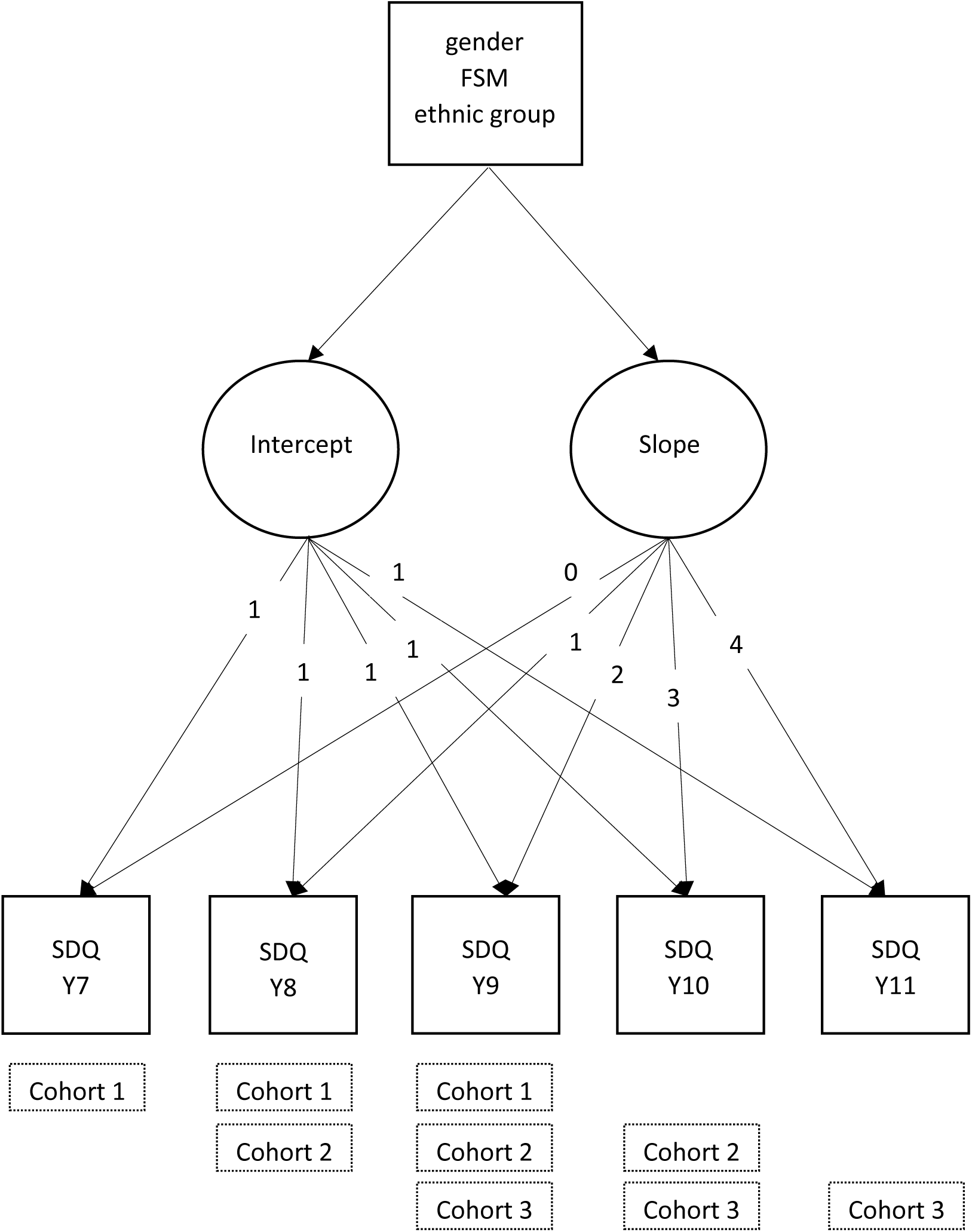
Latent growth curve model – path diagram. Abbreviations: Y7, school year 7; Y8, school year 8; Y9, school year 9; Y10, school year 10; Y11, school year 11; FSM, free school meals status; SDQ, strengths and difficulties questionnaire scores, representing three distress metrics, modelled separately, i.e., total difficulties scores, internalising scores, and externalising scores. (Note: residual covariances and covariance of the intercept and slope are omitted from diagram but present in substantive model.)

### Young persons’ perspectives

This paper is co-written with five young people from inner-London schools (co-authors TR, AH, KSCG, JK and NT) who work on REACH as young co-researchers. Their input is woven throughout the paper and shaped the hypotheses, interpretation, discussion, and conclusions. Their reflections on this work are provided in their own words in Supporting Information Appendix S2.

## RESULTS

### Sample characteristics

Of 4945 eligible students, 4663 (94.3%) provided valid SDQ data at one or more waves and are included in this analysis (Table 1). Of those included, 3034 (65%) provided SDQ data at all three waves, 1160 (25%) at two waves, and 469 (10%) at one wave. Each cohort (C1-C3) comprises around a third of the analysis sample (C1: n, 1690, 36.2%; C2: n, 1511, 32.4%; C3: n, 1462, 31.4%). Those included are highly representative of the target population based on key demographic characteristics (Table 1) (Knowles et al., 2022), with over a quarter receiving free school meals (n, 1321, 28.7%), half girls (n, 2355, 50.5%), and over 80% from minoritised ethnic groups. The largest ethnic groups are Black African (n, 1195, 25.6%), Black Caribbean (n, 764, 16.4%), White British (n, 686, 14.7%), and Mixed Black-and-White (n, 426, 9.1%).

**Table 1.** Sample characteristics.

|  | All<br>(n, 4663) |  | Cohort* |  |  |  |  |  | Gender |  |  |  | Free school meals |  |  |  |
| --- | --- | --- | --- | --- | --- | --- | --- | --- | --- | --- | --- | --- | --- | --- | --- | --- |
|  |  |  | Cohort 1<br>(n, 1690) |  | Cohort 2<br>(n, 1511) |  | Cohort 3<br>(n, 1462) |  | Boys<br>(n, 2308) |  | Girls<br>(n, 2355) |  | No<br>(n, 3277) |  | Yes<br>(n, 1321) |  |
|  | n | % | n | % | n | % | n | % | n | % | n | % | n | % | n | % |
| **Cohort** |  |  |  |  |  |  |  |  |  |  |  |  |  |  |  |  |
| 1 | 1690 | 36.2 | - | - | - | - | - | - | 835 | 36.2 | 855 | 36.3 | 1191 | 36.3 | 480 | 36.3 |
| 2 | 1511 | 32.4 | - | - | - | - | - | - | 753 | 32.6 | 758 | 32.2 | 1051 | 32.1 | 434 | 32.9 |
| 3 | 1462 | 31.4 | - | - | - | - | - | - | 720 | 31.2 | 742 | 31.5 | 1035 | 31.6 | 407 | 30.8 |
| **Gender** |  |  |  |  |  |  |  |  |  |  |  |  |  |  |  |  |
| Boy | 2308 | 49.5 | 835 | 49.4 | 753 | 49.8 | 720 | 49.3 | - | - | - | - | 1620 | 49.4 | 652 | 49.4 |
| Girl | 2355 | 50.5 | 855 | 50.6 | 758 | 50.2 | 742 | 50.8 | - | - | - | - | 1657 | 50.6 | 669 | 50.6 |
| **Free school meals** |  |  |  |  |  |  |  |  |  |  |  |  |  |  |  |  |
| No | 3277 | 71.3 | 1191 | 71.3 | 1051 | 70.8 | 1035 | 71.8 | 1620 | 71.3 | 1657 | 71.2 | - | - | - | - |
| Yes | 1321 | 28.7 | 480 | 28.7 | 434 | 29.2 | 407 | 28.2 | 652 | 28.7 | 669 | 28.8 | - | - | - | - |
| **Ethnic group** |  |  |  |  |  |  |  |  |  |  |  |  |  |  |  |  |
| Black African | 1195 | 25.6 | 408 | 24.1 | 398 | 26.3 | 389 | 26.6 | 587 | 25.4 | 608 | 25.8 | 845 | 25.8 | 335 | 25.4 |
| Black Caribbean | 764 | 16.4 | 241 | 14.3 | 274 | 18.1 | 249 | 17.0 | 375 | 16.3 | 389 | 16.5 | 475 | 14.5 | 277 | 21.0 |
| White British | 686 | 14.7 | 290 | 17.2 | 187 | 12.4 | 209 | 14.3 | 352 | 15.3 | 334 | 14.2 | 550 | 16.8 | 131 | 9.9 |
| Mixed Black and White | 426 | 9.1 | 159 | 9.4 | 134 | 8.9 | 133 | 9.1 | 207 | 9.0 | 219 | 9.3 | 263 | 8.0 | 158 | 12.0 |
| Other | 1592 | 34.1 | 592 | 35.0 | 518 | 34.3 | 482 | 33.0 | 787 | 34.1 | 805 | 34.2 | 1144 | 34.9 | 420 | 31.8 |

\*Cohort 1: age 11-12 years (school year 7) at baseline; Cohort 2: age 12-13 years (school year 8) at baseline; Cohort 3: age 13-14 years (school year 9) at baseline. See Knowles (2022) for sociodemographic characteristics of the target population.

### Repeated measures of mental distress

Summary statistics and correlation matrices for the repeated measures of distress are presented in Table 2. As expected, distress scores were strongly positively correlated over time, with correlation coefficients ranging from around 0.5 to 0.7. Total difficulties scores were approximately normally distributed within each school year (Figure S2). Internalising and externalising scores were mildly skewed, but within the accepted range of skewness and kurtosis for LCM with robust estimation.

**Table 2.** Correlation matrix and summary statistics for the repeated measures of mental distress.

| Correlation matrix |  |  |  |  |  | Summary statistics |  |  |
| --- | --- | --- | --- | --- | --- | --- | --- | --- |
| Total difficulties scores |  |  |  |  |  |  |  |  |
|  | year 7 | year 8 | year 9 | year 10 | year 11 | n | mean | sd |
| year 7 | 1.000 |  |  |  |  | 1560 | 12.2 | 6.0 |
| year 8 | 0.619 | 1.000 |  |  |  | 2837 | 12.0 | 5.8 |
| year 9 | 0.569 | 0.669 | 1.000 |  |  | 3971 | 12.0 | 5.8 |
| year 10 | . | 0.528 | 0.675 | 1.000 |  | 2326 | 11.8 | 5.7 |
| year 11 | . | . | 0.579 | 0.725 | 1.000 | 934 | 11.8 | 5.8 |
| Internalising scores |  |  |  |  |  |  |  |  |
|  | year 7 | year 8 | year 9 | year 10 | year 11 | n | mean | sd |
| year 7 | 1.000 |  |  |  |  | 1577 | 5.6 | 3.5 |
| year 8 | 0.568 | 1.000 |  |  |  | 2858 | 5.5 | 3.5 |
| year 9 | 0.496 | 0.638 | 1.000 |  |  | 4002 | 5.6 | 3.5 |
| year 10 | . | 0.535 | 0.654 | 1.000 |  | 2341 | 5.7 | 3.4 |
| year 11 | . | . | 0.543 | 0.716 | 1.000 | 939 | 5.8 | 3.5 |
| Externalising scores |  |  |  |  |  |  |  |  |
|  | year 7 | year 8 | year 9 | year 10 | year 11 | n | mean | sd |
| year 7 | 1.000 |  |  |  |  | 1576 | 6.5 | 3.8 |
| year 8 | 0.658 | 1.000 |  |  |  | 2863 | 6.5 | 3.7 |
| year 9 | 0.596 | 0.677 | 1.000 |  |  | 4003 | 6.5 | 3.6 |
| year 10 | . | 0.522 | 0.661 | 1.000 |  | 2341 | 6.2 | 3.4 |
| year 11 | . | . | 0.589 | 0.694 | 1.000 | 938 | 6.0 | 3.3 |

For all correlations, $p < 0.0001$ . Abbreviations: n, frequency; sd, standard deviation.

### Distress trajectories, by group Gender

There was no evidence of a difference in mean levels of general distress between boys and girls in Year 7 (difference in mean intercepts: 0.20 [95% CI -0.28, 0.67]), but strong evidence of a difference in mean rate of change from Year 7 to 11 (difference in slopes: 0.71 [0.54, 0.88]) (Table 3 and Figure 2). On average, distress increased with age among girls and decreased with age among boys. Thus, consistent with our hypothesis, the gap between boys and girls widened year-on-year (Figure 2). This pattern was entirely driven by internalising scores, for which the difference, though modest, was already evident in Year 7 (0.73 [0.51, 0.95]). For externalising trajectories, there was strong evidence of higher levels among boys (vs. girls) in Year 7 (−0.55 [-0.87, -0.23]), but the mean slopes converged over time due to decreases with age, on average, among boys (slope: -0.26 [-0.35, -0.16]) and stability among girls. Adjusting for FSM and ethnic group made no differences to these estimates (Table 3).

**Table 3.**
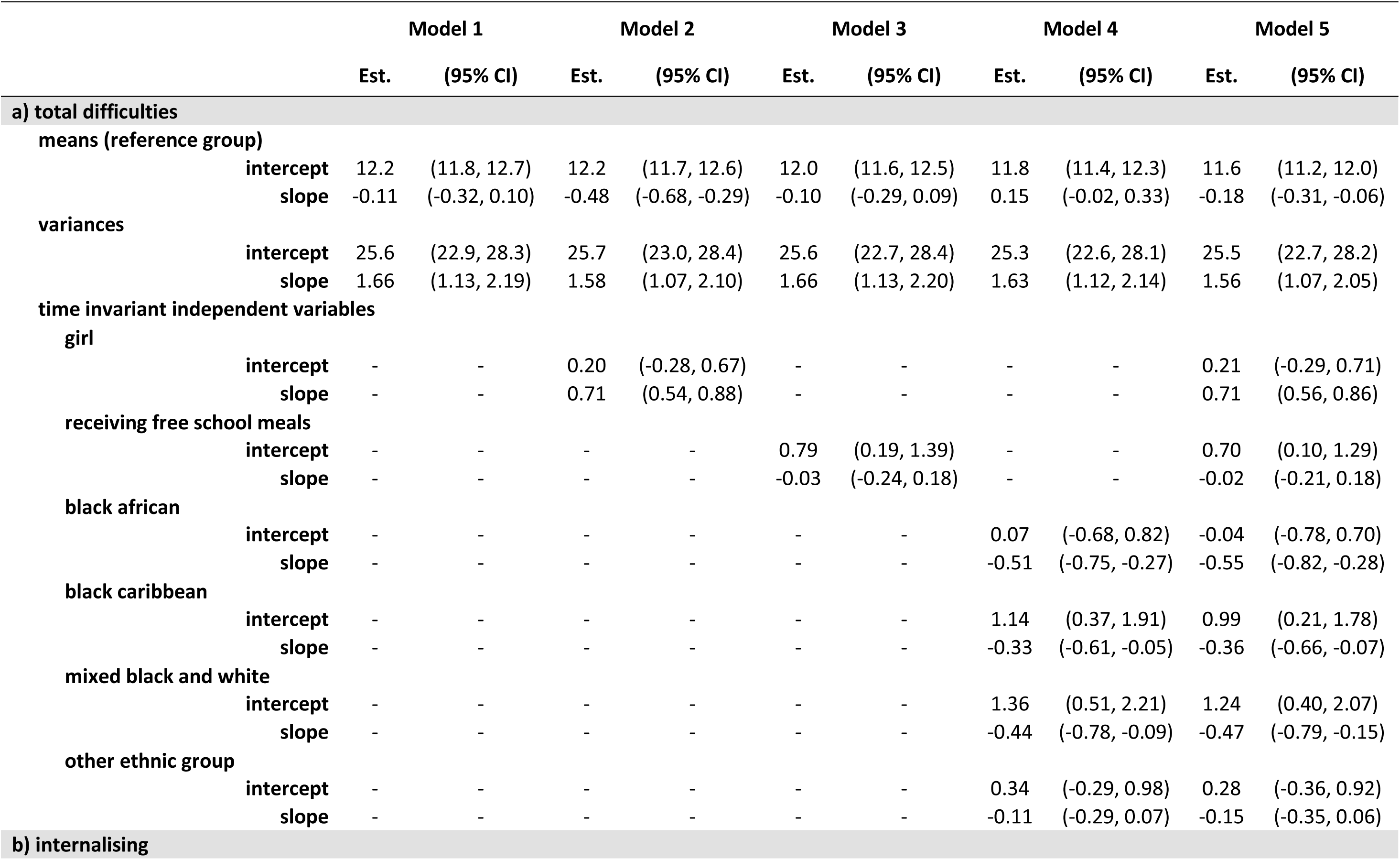
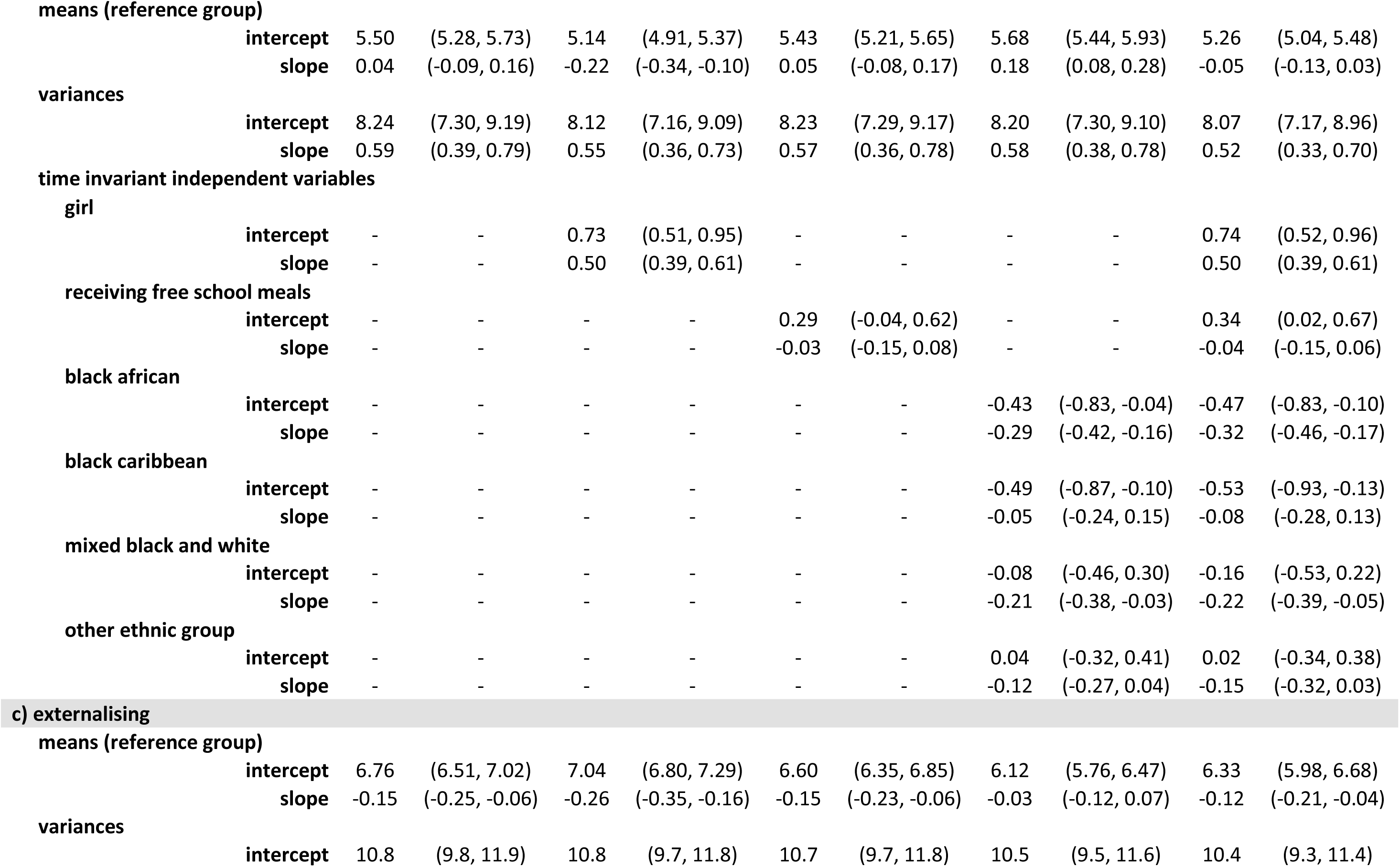
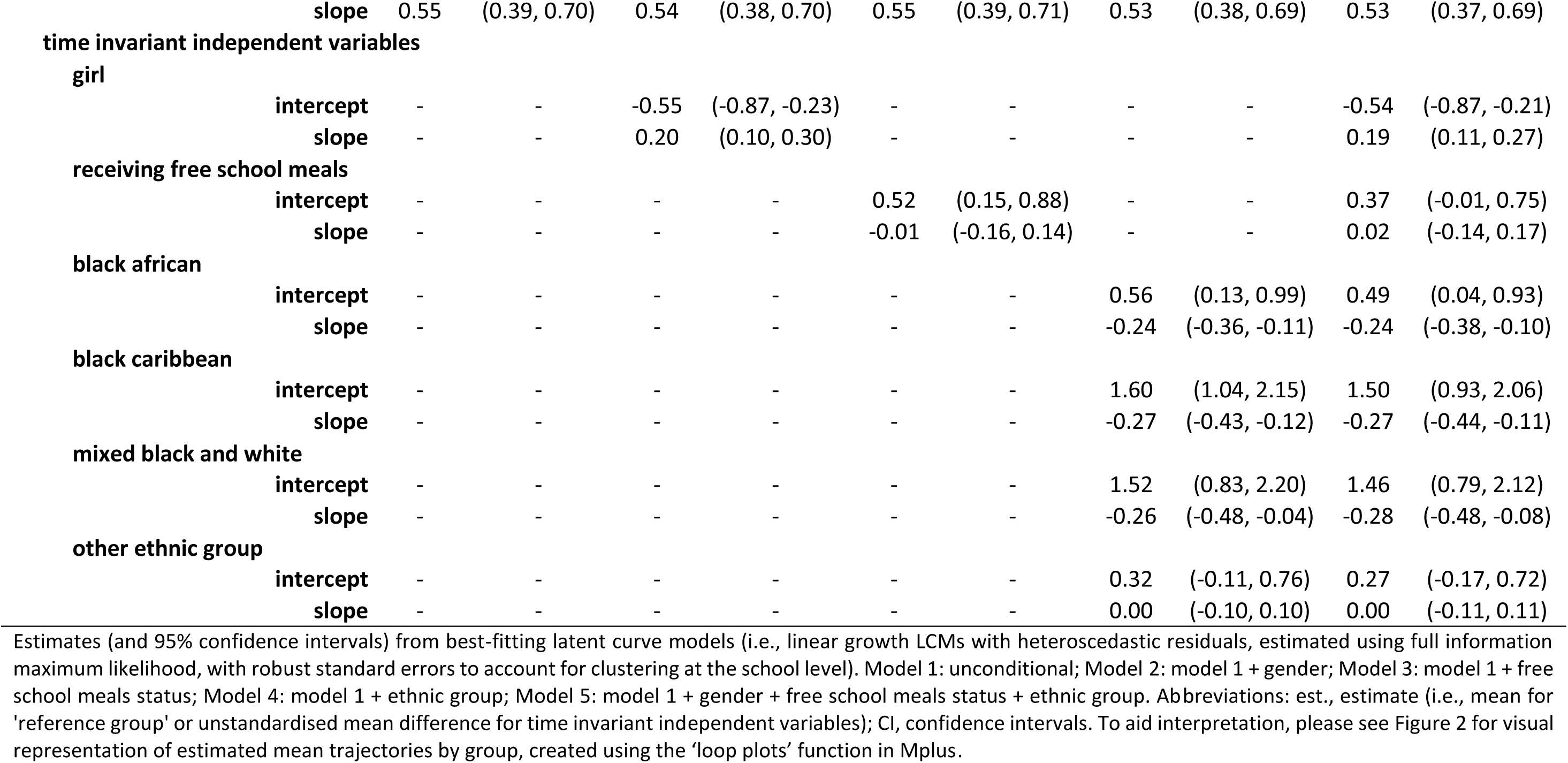
Growth parameter estimates from best-fitting latent growth curve model.

|  |  | Model 1 |  | Model 2 |  | Model 3 |  | Model 4 |  | Model 5 |  |
| --- | --- | --- | --- | --- | --- | --- | --- | --- | --- | --- | --- |
|  |  | Est. | (95% CI) | Est. | (95% CI) | Est. | (95% CI) | Est. | (95% CI) | Est. | (95% CI) |
| **a) total difficulties** |  |  |  |  |  |  |  |  |  |  |  |
| **means (reference group)** |  |  |  |  |  |  |  |  |  |  |  |
|  | **intercept** | 12.2 | (11.8, 12.7) | 12.2 | (11.7, 12.6) | 12.0 | (11.6, 12.5) | 11.8 | (11.4, 12.3) | 11.6 | (11.2, 12.0) |
|  | **slope** | -0.11 | (-0.32, 0.10) | -0.48 | (-0.68, -0.29) | -0.10 | (-0.29, 0.09) | 0.15 | (-0.02, 0.33) | -0.18 | (-0.31, -0.06) |
| **variances** |  |  |  |  |  |  |  |  |  |  |  |
|  | **intercept** | 25.6 | (22.9, 28.3) | 25.7 | (23.0, 28.4) | 25.6 | (22.7, 28.4) | 25.3 | (22.6, 28.1) | 25.5 | (22.7, 28.2) |
|  | **slope** | 1.66 | (1.13, 2.19) | 1.58 | (1.07, 2.10) | 1.66 | (1.13, 2.20) | 1.63 | (1.12, 2.14) | 1.56 | (1.07, 2.05) |
| **time invariant independent variables** |  |  |  |  |  |  |  |  |  |  |  |
| **girl** |  |  |  |  |  |  |  |  |  |  |  |
|  | **intercept** | - | - | 0.20 | (-0.28, 0.67) | - | - | - | - | 0.21 | (-0.29, 0.71) |
|  | **slope** | - | - | 0.71 | (0.54, 0.88) | - | - | - | - | 0.71 | (0.56, 0.86) |
| **receiving free school meals** |  |  |  |  |  |  |  |  |  |  |  |
|  | **intercept** | - | - | - | - | 0.79 | (0.19, 1.39) | - | - | 0.70 | (0.10, 1.29) |
|  | **slope** | - | - | - | - | -0.03 | (-0.24, 0.18) | - | - | -0.02 | (-0.21, 0.18) |
| **black african** |  |  |  |  |  |  |  |  |  |  |  |
|  | **intercept** | - | - | - | - | - | - | 0.07 | (-0.68, 0.82) | -0.04 | (-0.78, 0.70) |
|  | **slope** | - | - | - | - | - | - | -0.51 | (-0.75, -0.27) | -0.55 | (-0.82, -0.28) |
| **black caribbean** |  |  |  |  |  |  |  |  |  |  |  |
|  | **intercept** | - | - | - | - | - | - | 1.14 | (0.37, 1.91) | 0.99 | (0.21, 1.78) |
|  | **slope** | - | - | - | - | - | - | -0.33 | (-0.61, -0.05) | -0.36 | (-0.66, -0.07) |
| **mixed black and white** |  |  |  |  |  |  |  |  |  |  |  |
|  | **intercept** | - | - | - | - | - | - | 1.36 | (0.51, 2.21) | 1.24 | (0.40, 2.07) |
|  | **slope** | - | - | - | - | - | - | -0.44 | (-0.78, -0.09) | -0.47 | (-0.79, -0.15) |
| **other ethnic group** |  |  |  |  |  |  |  |  |  |  |  |
|  | **intercept** | - | - | - | - | - | - | 0.34 | (-0.29, 0.98) | 0.28 | (-0.36, 0.92) |
|  | **slope** | - | - | - | - | - | - | -0.11 | (-0.29, 0.07) | -0.15 | (-0.35, 0.06) |
| **b) internalising** |  |  |  |  |  |  |  |  |  |  |  |

**means (reference group)**
|  |  |  |  |  |  |  |  |  |  |  |
| --- | --- | --- | --- | --- | --- | --- | --- | --- | --- | --- |
| **intercept** | 5.50 | (5.28, 5.73) | 5.14 | (4.91, 5.37) | 5.43 | (5.21, 5.65) | 5.68 | (5.44, 5.93) | 5.26 | (5.04, 5.48) |
| **slope** | 0.04 | (-0.09, 0.16) | -0.22 | (-0.34, -0.10) | 0.05 | (-0.08, 0.17) | 0.18 | (0.08, 0.28) | -0.05 | (-0.13, 0.03) |

**variances**
|  |  |  |  |  |  |  |  |  |  |  |
| --- | --- | --- | --- | --- | --- | --- | --- | --- | --- | --- |
| **intercept** | 8.24 | (7.30, 9.19) | 8.12 | (7.16, 9.09) | 8.23 | (7.29, 9.17) | 8.20 | (7.30, 9.10) | 8.07 | (7.17, 8.96) |
| **slope** | 0.59 | (0.39, 0.79) | 0.55 | (0.36, 0.73) | 0.57 | (0.36, 0.78) | 0.58 | (0.38, 0.78) | 0.52 | (0.33, 0.70) |

**girl**
|  |  |  |  |  |  |  |  |  |  |  |
| --- | --- | --- | --- | --- | --- | --- | --- | --- | --- | --- |
| **intercept** | - | - | 0.73 | (0.51, 0.95) | - | - | - | - | 0.74 | (0.52, 0.96) |
| **slope** | - | - | 0.50 | (0.39, 0.61) | - | - | - | - | 0.50 | (0.39, 0.61) |

**receiving free school meals**
|  |  |  |  |  |  |  |  |  |  |  |
| --- | --- | --- | --- | --- | --- | --- | --- | --- | --- | --- |
| **intercept** | - | - | - | - | 0.29 | (-0.04, 0.62) | - | - | 0.34 | (0.02, 0.67) |
| **slope** | - | - | - | - | -0.03 | (-0.15, 0.08) | - | - | -0.04 | (-0.15, 0.06) |

**black african**
|  |  |  |  |  |  |  |  |  |  |  |
| --- | --- | --- | --- | --- | --- | --- | --- | --- | --- | --- |
| **intercept** | - | - | - | - | - | - | -0.43 | (-0.83, -0.04) | -0.47 | (-0.83, -0.10) |
| **slope** | - | - | - | - | - | - | -0.29 | (-0.42, -0.16) | -0.32 | (-0.46, -0.17) |

**black caribbean**
|  |  |  |  |  |  |  |  |  |  |  |
| --- | --- | --- | --- | --- | --- | --- | --- | --- | --- | --- |
| **intercept** | - | - | - | - | - | - | -0.49 | (-0.87, -0.10) | -0.53 | (-0.93, -0.13) |
| **slope** | - | - | - | - | - | - | -0.05 | (-0.24, 0.15) | -0.08 | (-0.28, 0.13) |

**mixed black and white**
|  |  |  |  |  |  |  |  |  |  |  |
| --- | --- | --- | --- | --- | --- | --- | --- | --- | --- | --- |
| **intercept** | - | - | - | - | - | - | -0.08 | (-0.46, 0.30) | -0.16 | (-0.53, 0.22) |
| **slope** | - | - | - | - | - | - | -0.21 | (-0.38, -0.03) | -0.22 | (-0.39, -0.05) |

**other ethnic group**
|  |  |  |  |  |  |  |  |  |  |  |
| --- | --- | --- | --- | --- | --- | --- | --- | --- | --- | --- |
| **intercept** | - | - | - | - | - | - | 0.04 | (-0.32, 0.41) | 0.02 | (-0.34, 0.38) |
| **slope** | - | - | - | - | - | - | -0.12 | (-0.27, 0.04) | -0.15 | (-0.32, 0.03) |

**means (reference group)**
|  |  |  |  |  |  |  |  |  |  |  |
| --- | --- | --- | --- | --- | --- | --- | --- | --- | --- | --- |
| **intercept** | 6.76 | (6.51, 7.02) | 7.04 | (6.80, 7.29) | 6.60 | (6.35, 6.85) | 6.12 | (5.76, 6.47) | 6.33 | (5.98, 6.68) |
| **slope** | -0.15 | (-0.25, -0.06) | -0.26 | (-0.35, -0.16) | -0.15 | (-0.23, -0.06) | -0.03 | (-0.12, 0.07) | -0.12 | (-0.21, -0.04) |

**variances**
|  |  |  |  |  |  |  |  |  |  |  |
| --- | --- | --- | --- | --- | --- | --- | --- | --- | --- | --- |
| **intercept** | 10.8 | (9.8, 11.9) | 10.8 | (9.7, 11.8) | 10.7 | (9.7, 11.8) | 10.5 | (9.5, 11.6) | 10.4 | (9.3, 11.4) |
| **slope** |  | 0.55 | (0.39, 0.70) | 0.54 | (0.38, 0.70) | 0.55 | (0.39, 0.71) | 0.53 | (0.38, 0.69) | 0.53 | (0.37, 0.69) |
| **time invariant independent variables** |  |  |  |  |  |  |  |  |  |  |  |
| **girl** |  |  |  |  |  |  |  |  |  |  |  |
|  | **intercept** | - | - | -0.55 | (-0.87, -0.23) | - | - | - | - | -0.54 | (-0.87, -0.21) |
|  | **slope** | - | - | 0.20 | (0.10, 0.30) | - | - | - | - | 0.19 | (0.11, 0.27) |
| **receiving free school meals** |  |  |  |  |  |  |  |  |  |  |  |
|  | **intercept** | - | - | - | - | 0.52 | (0.15, 0.88) | - | - | 0.37 | (-0.01, 0.75) |
|  | **slope** | - | - | - | - | -0.01 | (-0.16, 0.14) | - | - | 0.02 | (-0.14, 0.17) |
| **black african** |  |  |  |  |  |  |  |  |  |  |  |
|  | **intercept** | - | - | - | - | - | - | 0.56 | (0.13, 0.99) | 0.49 | (0.04, 0.93) |
|  | **slope** | - | - | - | - | - | - | -0.24 | (-0.36, -0.11) | -0.24 | (-0.38, -0.10) |
| **black caribbean** |  |  |  |  |  |  |  |  |  |  |  |
|  | **intercept** | - | - | - | - | - | - | 1.60 | (1.04, 2.15) | 1.50 | (0.93, 2.06) |
|  | **slope** | - | - | - | - | - | - | -0.27 | (-0.43, -0.12) | -0.27 | (-0.44, -0.11) |
| **mixed black and white** |  |  |  |  |  |  |  |  |  |  |  |
|  | **intercept** | - | - | - | - | - | - | 1.52 | (0.83, 2.20) | 1.46 | (0.79, 2.12) |
|  | **slope** | - | - | - | - | - | - | -0.26 | (-0.48, -0.04) | -0.28 | (-0.48, -0.08) |
| **other ethnic group** |  |  |  |  |  |  |  |  |  |  |  |
|  | **intercept** | - | - | - | - | - | - | 0.32 | (-0.11, 0.76) | 0.27 | (-0.17, 0.72) |
|  | **slope** | - | - | - | - | - | - | 0.00 | (-0.10, 0.10) | 0.00 | (-0.11, 0.11) |

**Figure 2.**
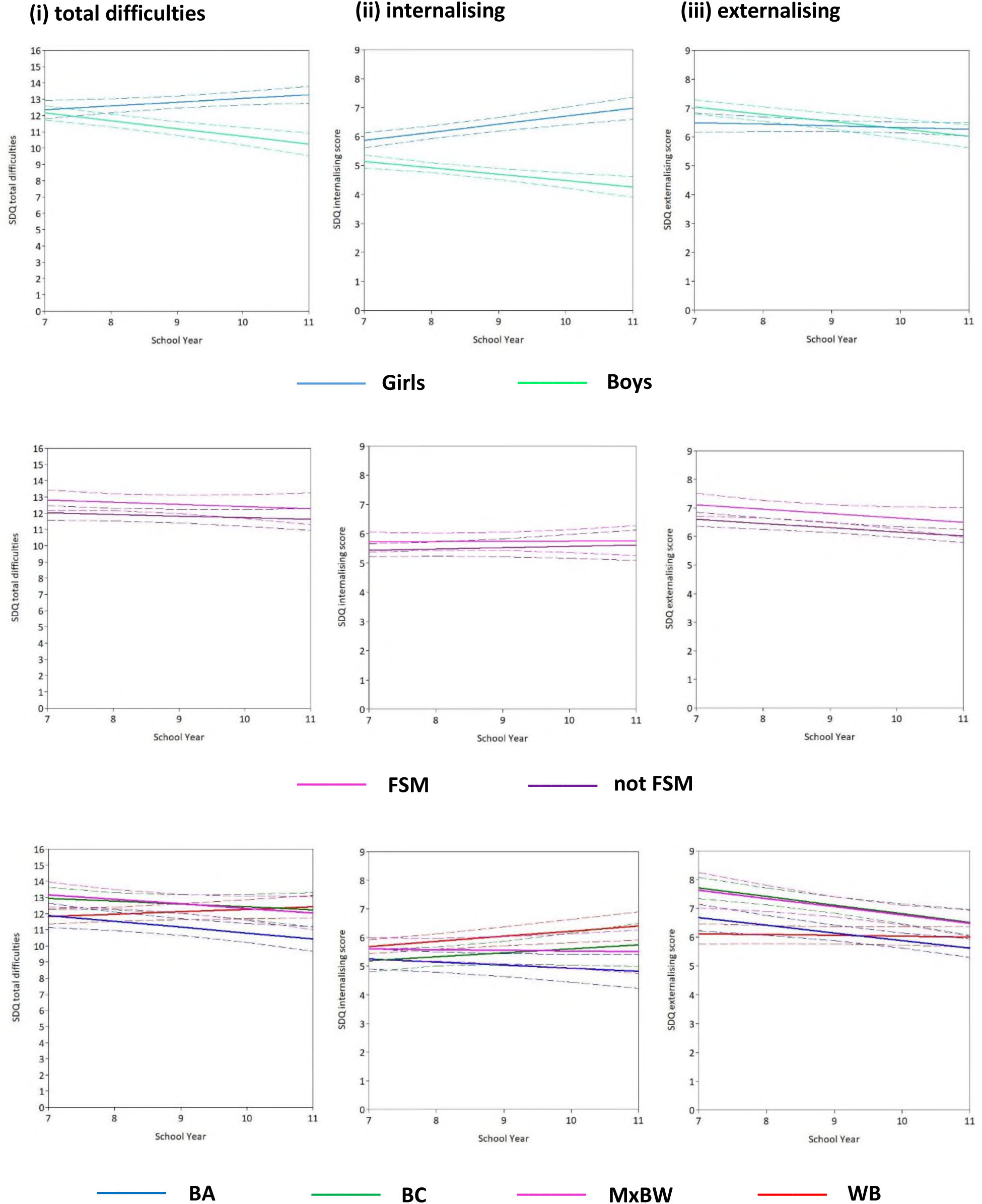
Mental health trajectories, by (a) gender, (b) free school meals, and (c) ethnic group. Solid lines: estimated mean trajectory per group. Dashed lines: 95% confidence intervals. Abbreviations: FSM, free school meals; BA, Black African; BC, Black Caribbean; MxBW, Mixed Black-and-White; WB, White British. Note: ‘Other’ ethnic group omitted due to limited interpretability (see Discussion), but parameter estimates are provided in Table 3. Unadjusted models are presented. See Table 3 for adjusted (and unadjusted) parameter estimates. Model fit statistics are provided in Supplementary Table S1.

### Free school meals

Consistent with our hypothesis, there was strong evidence of higher mean starting levels of general distress among those receiving (vs. not receiving) FSM (0.79 [95% CI: 0.19, 1.39]) but no evidence of a difference in rate of change between the two groups (−0.03 [-0.15, 0.18]) (Table 3, Figure 2). Broadly, this pattern was evident for both internalising and externalising scores; the difference in mean intercepts was slightly larger for externalising scores than for internalising scores in unadjusted models, but similar in adjusted models (Table 3).

### Ethnic group

Overall, we found mixed and dynamic patterns by ethnic group (Figure 2). For externalising trajectories, there was strong evidence of variability by ethnic group in mean intercepts and slopes and, broadly, these patterns mirrored those observed for general distress. Specifically, initial levels (intercepts) were highest among Black Caribbean and Mixed Black-and-White groups – who shared a very similar mean trajectory – and lowest among White British (e.g., difference in intercepts Black Caribbean vs White British: 1.60 [1.04, 2.15]). Thereafter, the slope was stable on average among White British (mean slope: -0.03 [-.012, 0.07]) but decreased, on average, among Black African (difference: -0.24 [-0.36, -0.11]), Black Caribbean (difference: -0.27 [-0.43, -0.12]), and Mixed Black-and-White (difference: -0.26 [-0.48, -0.04]) groups. As such, the initial differences narrowed with age. By Year 11, Black African youth reported the lowest levels of externalising distress, on average.

For internalising scores, there was some evidence of variability in mean intercepts by ethnic group, but the differences were modest, e.g., mean starting level was around 0.5 points lower among Black African and Black Caribbean compared with White British and Mixed Black-and-White groups (e.g., Black Caribbean vs. White British: -0.49 [-0.87, -0.10]). Thereafter, internalising slopes increased at similar rates on average among White British and Black Caribbean, decreased on average among Black African, and remained stable among Mixed Black-and-White groups. As such, by mid-to-late secondary school Black African youth fared better than all other groups and the White British group fared worse. Adjusting for gender and FSM made no substantive difference to these estimates (Table 3).

### Distress trajectories, by intersecting identities

In some areas, our hypothesis was broadly supported but, overall, the results were complex, dependent on distress metric, age, and combination of identities. We outline here some broad themes, but direct the reader to Figure 3, Figure S3, and Table S2 for full results.

**Figure 3.**
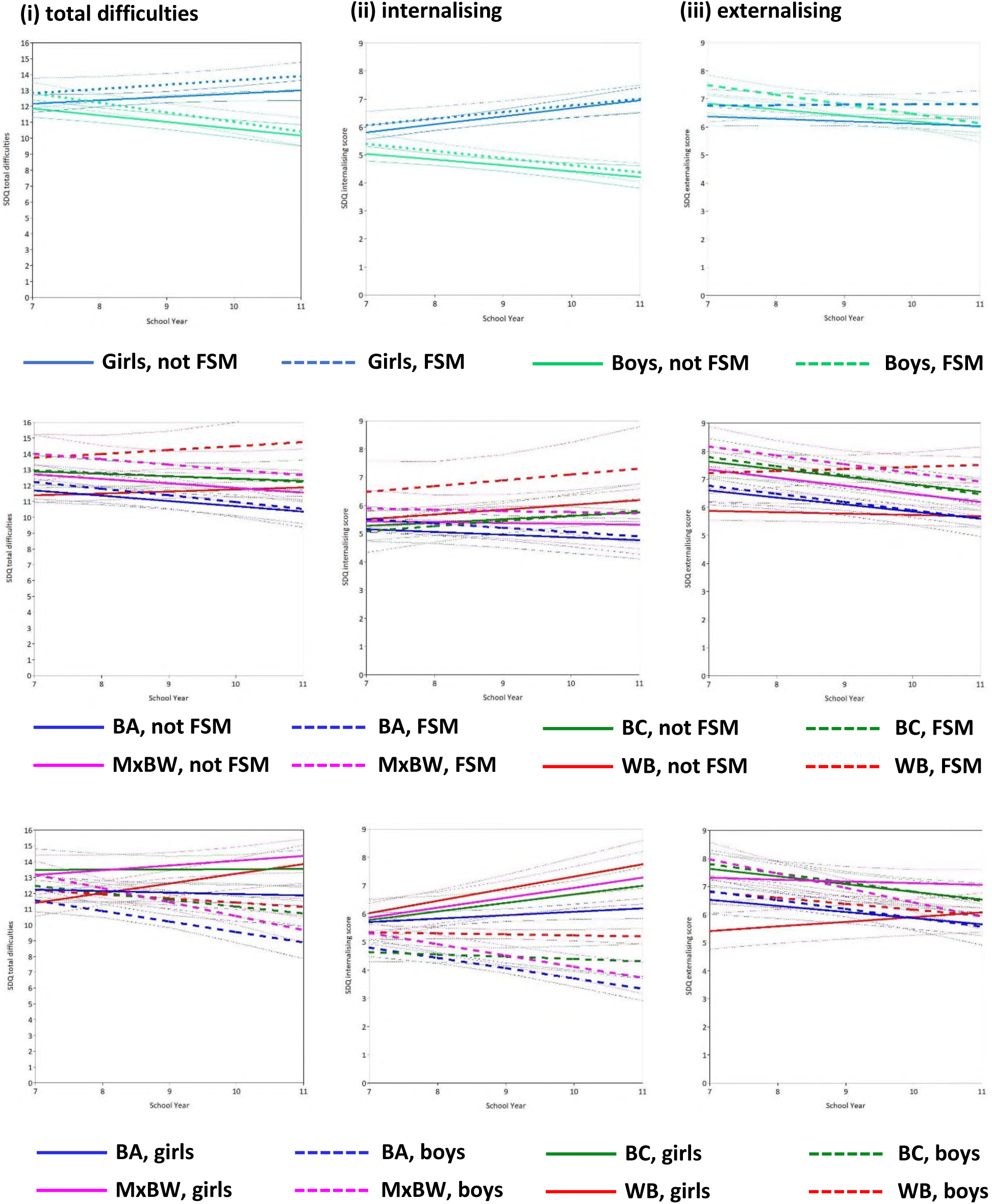
Mental health trajectories, by (crude) intersecting identities: (a) gender*FSM, (b) ethnic group*FSM, (c) ethnic group*gender. Bold lines: estimated mean trajectory per group. Faint dashed lines: 95% confidence intervals. Abbreviations: FSM, free school meals; BA, Black African; BC, Black Caribbean; MxBW, Mixed Black-and-White; WB, White British. Note: ‘Other’ ethnic group omitted due to limited interpretability (see Discussion), but parameter estimates are provided in Table S2. Unadjusted models are presented. See Supplementary Table S2 for adjusted (and unadjusted) parameter estimates.

First, there was very little evidence of interactions between gender and FSM in relation to mean intercepts or slopes for any distress metric (Table S2). That is, gender differences in distress trajectories were consistent across strata of FSM, and vice versa (Figure 3). Second, there was strong evidence of interactions between ethnic group and FSM, and between ethnic group and gender, and broadly these related to externalising, not internalising, scores (Table S2). That is, the magnitude of inequalities in distress – particularly externalising – trajectories by gender and by FSM varied by ethnic group. Specifically, for externalising trajectories, there was strong evidence of higher starting levels among boys (vs. girls) and those receiving FSM (vs. not) within the White British and Mixed Black-and-White groups, but little-to-no evidence of such differences within the Black African and Black Caribbean groups (Figure 3, Table S2). Third, across all distress metrics, the Black African group seemed to do relatively well throughout secondary school compared with their peers, and this was the case overall and within strata of gender and FSM. In relation to interactions between FSM and ethnic group *within* gender (Figure S3), White British girls on FSM – and, to some extent, Mixed Black-and-White girls on FSM – generally seemed to fare worse than other groups by the end of secondary school. By Year 11, White British girls on FSM had the highest internalising levels of all groups.

## DISCUSSION

### Key findings

To our knowledge, this study is the first in the UK to examine inequalities in trajectories of mental distress in young people from diverse inner-city communities. Our work strengthens, extends, and introduces new evidence to the field in several ways. First, our findings strongly suggest that gender inequalities in internalising distress emerge in late childhood/early adolescence and widen year-on-year through adolescence. Broadly, this pattern was evident in all ethnic groups and in lower and higher income groups. Second, we found strong evidence of higher initial levels of distress, on average, among young people from lower (vs. higher) income households, and this gap, though modest in some groups, persisted throughout secondary school. Third, by ethnic group and intersecting identities, the picture was somewhat complex (e.g., dependent on combination of identities), dynamic (i.e., changed with age) and mixed (i.e., varied by type of distress). Overall and on average, young people from Black African backgrounds generally reported better mental health through much of secondary school compared with their peers; those of Black Caribbean and Mixed Black-and-White backgrounds shared similar trajectories, differing somewhat from those of Black African background; and by age 16, lower-income White British girls reported the highest levels of internalising distress.

### Limitations

Our work has several notable strengths, including the large, diverse, representative cohorts; the longitudinal and accelerated design; the relatively short intervals between waves; the use of self-report data; the focus on the period of the lifecourse in which many mental health problems first emerge (Kessler et al., 2007; Jones, 2013); and coproduction with young people. These strengths address many of the limitations of existing evidence on the developmental origins of social inequalities in mental health. Nonetheless, several important limitations should be noted. First, as with all measures of adolescent mental health, the SDQ is imperfect, and it is possible that some groups systematically under-report distress. Second, our measure of household income is crude and limited. It does not capture the full range of incomes and while most who receive FSM will indeed be from low-income households, many who are not eligible will nonetheless be experiencing financial hardship. Indeed, by recent estimates, over a third of children (around 900,000) living in poverty are not eligible for FSM (Patrick et al., 2021). However, FSM is widely used in school-based studies because younger students tend not to be able to provide information on alternative indicators of income/SES (e.g., parental income, occupation), whereas almost all can accurately report their FSM status. Third, our approach to exploring ‘intersectionality’ is basic and limited. Methods for modelling intersectionality in large population-based studies are emerging (Bauer et al., 2021) but are yet to be extended to more complex longitudinal data structures. We monitor their development with interest.

### Findings in context

Prior to REACH, there was a distinct lack of robust and representative data on the mental health of young people from diverse backgrounds in the UK. Where data does exist, it is generally nationally representative, cross-sectional, predominantly White, under-represents marginalised groups, permits mostly broad and single-identity groupings, and/or relates to prior generations (Clark et al., 2007; Maynard et al., 2007; McElroy et al., 2023; Sadler et al., 2018; Smith et al., 2015). Overall, these studies suggest a ‘mental health advantage’ among young people from minoritised ethnic groups (Bhui et al., 2012; Goodman et al., 2008; Harding et al., 2015; Sadler et al., 2018), but the evidence is limited and mixed. Only one other study, the Millennium Cohort Study (MCS), provides information on the extent and nature of racial/ethnic inequalities in mental health trajectories in contemporary youth (Bains & Gutman, 2021; Gutman, 2019; Terhaag et al., 2021; Zilanawala et al., 2018). Broadly, analyses of (parent-report) MCS data suggest: (a) for internalising scores, most minoritised ethnic groups fare worse than their white peers during *childhood* (age 3-5/7), but for some groups (e.g. Black African, Indian) this reverses in early adolescence, such that Black African and Indian youth generally fare better whereas all other groups seem to converge to relatively similar levels by age 14 (Bains & Gutman, 2021); (b) for externalising scores, young people from Black African backgrounds generally fare better than other groups in childhood and early adolescence (Bains & Gutman, 2021); and (c) in some (not all) groups, those of ‘mixed’ ethnicity fare worse on externalising scores compared with their ‘non-mixed’ peers (Zilanawala et al., 2018). Our findings largely support their observations of relatively good (reported) mental health trajectories among young people from Black African backgrounds; of clear differences in trajectories between Black African and Black Caribbean groups (the latter tending to fare worse); and of some interesting similarities by ethnic group despite different circumstances and experiences. Some of these patterns mirror those observed in adults, others less so. For instance, observations of lower reported distress among Black African youth are seemingly at odds with consistent evidence in adults of elevated rates of severe mental illness among Black African and Black Caribbean communities (vs. ‘White’, variously defined) (Morgan et al., 2006; Morgan et al., 2019), which may be associated with current, intergenerational, and historic social contexts (i.e., of racism and discrimination, social and economic oppression, exposure to violence, etc.) (Morgan et al., 2019; Nazroo et al., 2020). A better understanding of the contemporary experiences of young people from diverse racial and ethnic backgrounds and how these relate to the unfolding of inequalities in distress across the lifecourse is important for prevention strategies.

Our findings support a growing body of evidence that places the onset and worsening of gender inequalities in emotional/internalising problems in late childhood/early adolescence (Clark et al., 2007; Terhaag et al., 2021; Yoon et al., 2022). Further, our work strongly suggests this to be the case in higher and lower income groups and across diverse ethnic groups. Early adolescence may, then, be the period in which lifelong gender inequalities in mental health could be mitigated or prevented. However, our understanding of the causes of gender inequalities in mental distress is remarkably limited (Patalay & Demkowicz, 2023; Kuehner, 2017; Piccinelli & Wilkinson, 2000). In adults, there is growing evidence to support a role of systemic gender inequity, e.g., as manifests in things like unequal pay, unequal burden of unpaid work, and gender-based violence (Fergusson et al., 2002; Platt et al., 2016; Xue & McMunn, 2021). Similarly, the ‘gender depression gap’ in mid-to-late adolescence may, in part, be explained by sexual violence (Bentivegna & Patalay, 2022). There is also evidence that the size of gender mental health gaps in mid-adolescence varies widely across countries, suggesting a role of the social environment (Campbell et al., 2021). The causes are likely complex and multifaceted, but there has been little attempt to systematically and comprehensively examine the dynamic interplay of possible causes – from the biological to the social, and issues of measurement – and thus how to intervene. This work is urgently needed.

Caveats notwithstanding, our findings are consistent with a robust body of evidence of inequalities in mental distress by SES/income, which are evident from an early age and persist over time (Allen et al., 2014; Terhaag et al., 2021; Wickham et al., 2017). For instance, there is strong evidence that children who transition into poverty experience a sharp increase in distress (Wickham et al., 2017) and that the odds of mental health problems in those who experience persistent poverty during childhood – by recent estimates, around one-in-five children in the UK – are around three times higher than in those who never experience poverty (Lai et al., 2019). Worryingly, there is also tentative evidence to suggest socioeconomic inequalities in mental health may be widening (Collishaw et al., 2019; McElroy et al., 2023) and the consequences of mental ill health worsening (Sellers et al., 2019) in newer generations. In a time of prolonged austerity, rising child poverty, and growing income and wealth inequality in the UK, there is a clear need for targeted support for low income and financially insecure households. Effective policies could greatly reduce inequalities in mental distress and, indeed, other outcomes (e.g., education) (Lai et al., 2019), with widespread and lasting benefits for individuals, families, and society.

### Implications

There is widespread concern about a ‘mental health crisis’ among young people in the UK. But there is also strong evidence, from our own and others’ work, of variation by social group in the levels and trajectories of mental distress among young people and in the extent to which levels and rates are rising. That is, the distribution of distress is socially structured. It follows that individual-focussed interventions (e.g., mindfulness training, talking therapies) (Ellins et al., 2023; Kuyken et al., 2022; Stallard et al., 2013) – which currently dominate the youth mental health policy agenda in the absence of any coherent prevention strategy (Department of Health & Department for Education, 2017) – are unlikely to be an effective solution to the problem. Indeed, there is emerging evidence that such interventions are of limited effectiveness, particularly for those with greatest need (Ellins et al., 2023; Kuyken et al., 2022; Montero-Marin et al., 2022). There is a real risk, then, that such interventions – and the policies that support their roll-out – could inadvertently perpetuate or exacerbate inequalities in mental health.

## Conclusion

In diverse inner-city communities, early-to-mid adolescence is an important period in the emergence and persistence of some of the social inequalities in mental health widely reported in adults; others are more nuanced and complex. Understanding the conditions and experiences that mitigate distress in disadvantaged and marginalised groups is an important next step.

## Key Points

- Our understanding of the age at which inequalities in mental distress emerge and how they evolve with age is limited.
- We used data from our diverse London-based youth cohorts, REACH, to examine trajectories of distress from age 11-16 years, overall and by gender, ethnic group, free school meals status (an indicator of household income), and their intersections.
- We found strong evidence of inequalities in trajectories of distress by gender and free school meals. By ethnic group and intersectional identities, the picture was more complex and dynamic, with interesting differences (e.g., between Black African and Black Caribbean youth) and similarities.
- The distribution of distress in adolescence is socially structured. Policies and approaches to support and promote youth mental health may benefit from greater consideration of the social drivers of distress.

## Data Availability

We welcome and encourage requests from those wishing to access REACH data for specific research projects or collaborations. Our data access policy, which aims to make REACH data as accessible as possible while adhering to legal and ethical principles and protecting the privacy of schools and participants, can be found at www.thereachstudy.com/information-for-researchers.html. Further information about REACH is also available on the study website. Applications should be submitted to the Principal Investigator, Professor Craig Morgan, at.

## ACKNOWLEDGMENTS

The authors would like to thank all participating schools, teachers, young people, and parents for taking part in and supporting REACH. The authors would also like to thank the research assistants and students who contributed to data collection.

## ETHICS APPROVAL

All study procedures were approved by the Psychiatry, Nursing and Midwifery Research Ethics Subcommittee (PNM-RESC), King’s College London (ref: 15/16-2320).

## SUPPLEMENTARY DATA

Supporting information is available online at [XXXXXXXXX].

## AUTHOR CONTRIBUTIONS

**GK**: collected and prepared data; coordinated REACH; planned and conducted the analyses; drafted and critically revised the manuscript; approved the final manuscript; **CGA**: collected and cleaned data; reviewed literature; verified the analyses/results; drafted and critically revised the manuscript; approved the final manuscript; **TR, AH, KSCG, JK, NT** (young co-researchers): informed research procedures and processes; shaped hypotheses; interpreted findings; co-wrote sections of the manuscript; critically revised the manuscript; approved the final manuscript; **SD, DS, AO, AT, EP, LD, RB, KL:** collected and cleaned data and/or mentored young co-researchers; critically revised the manuscript; approved the final manuscript; **VP, SH, KB: VP:** contributed to the conceptualisation and design of the work; interpreted findings; revised the manuscript; approved the final manuscript; **CM**: conceptualized, designed, and secured funding for REACH; collected data; supervised analyses; critically revised the manuscript; approved the final manuscript.

## DATA ACCESS

GK, CGA, and CM had full access to all the data and take responsibility for the integrity of the data and the accuracy of the analyses.

## Abbreviations

FSM: free school meals
SDQ: strengths and difficulties questionnaire
LCM: latent growth curve models

## REFERENCES

1. Allen, J., Balfour, R., Bell, R., & Marmot, M. (2014). Social determinants of mental health. International Review of Psychiatry, 26(4), 392–407.

2. Bains, S., & Gutman, L. M. (2021). Mental Health in Ethnic Minority Populations in the UK: Developmental Trajectories from Early Childhood to Mid Adolescence. Journal of Youth and Adolescence, 50(11), 2151–2165.

3. Bauer, G. R., Churchill, S. M., Mahendran, M., Walwyn, C., Lizotte, D., & Villa-Rueda, A. A. (2021). Intersectionality in quantitative research: A systematic review of its emergence and applications of theory and methods. SSM Population Health, 14, 100798. 10.1016/j.ssmph.2021.100798

4. Bentivegna, F., & Patalay, P. (2022). The impact of sexual violence in mid-adolescence on mental health: a UK population-based longitudinal study. Lancet Psychiatry, 9(11), 874–883.

5. Bhui, K., Stansfeld, S., Hull, S., Priebe, S., Mole, F., & Feder, G. (2003). Ethnic variations in pathways to and use of specialist mental health services in the UK: Systematic review. The British Journal of Psychiatry, 182(2), 105–116.

6. Bhui, K. S., Lenguerrand, E., Maynard, M. J., Stansfeld, S. A., & Harding, S. (2012). Does cultural integration explain a mental health advantage for adolescents? International Journal of Epidemiology, 41(3), 791–802.

7. Blakey, R., Morgan, C., Gayer-Anderson, C., Davis, S., Beards, S., Harding, S., Pinfold, V., Bhui, K., Knowles, G., & Viding, E. (2021). Prevalence of conduct problems and social risk factors in ethnically diverse inner-city schools. BMC Public Health, 21, 1–13.

8. Booth, C., Moreno-Agostino, D., & Fitzsimons, E. (2023). Parent-adolescent informant discrepancy on the Strengths and Difficulties Questionnaire in the UK Millennium Cohort Study. Child and Adolescent Psychiatry and Mental Health, 17(*1*), 57.

9. Campbell, O. L. K., Bann, D., & Patalay, P. (2021). The gender gap in adolescent mental health: A cross-national investigation of 566,829 adolescents across 73 countries. SSM Population Health, 13, 100742.

10. Clark, C., Haines, M. M., Head, J., Klineberg, E., Arephin, M., Viner, R., Taylor, S. J., Booy, R., Bhui, K., & Stansfeld, S. A. (2007). Psychological symptoms and physical health and health behaviours in adolescents: a prospective 2-year study in East London. Addiction, 102(1), 126–135.

11. Collishaw, S., Furzer, E., Thapar, A. K., & Sellers, R. (2019). Brief report: a comparison of child mental health inequalities in three UK population cohorts. European Child and Adolescent Psychiatry, 28(11), 1547–1549.

12. Crenshaw, K. (1989). Demarginalizing the Intersection of Race and Sex: A Black Feminist Critique of Antidiscrimination Doctrine, Feminist Theory and Antiracist Politics. University of Chicago Legal Forum, 1, 139–167.

13. Department of Health, & Department for Education. (2017). Transforming Children and Young People’s Mental Health Provision: Green Paper. https://assets.publishing.service.gov.uk/government/uploads/system/uploads/attachment_data/file/664855/Transforming_children_and_young_people_s_mental_health_provision.pdf. Last accessed: 5th May 2023.

14. Drosopoulou, G., Vlasopoulou, F., Panagouli, E., Stavridou, A., Papageorgiou, A., Psaltopoulou, T., Tsolia, M., Tzavara, C., Sergentanis, T. N., & Tsitsika, A. K. (2023). Cross-National Comparisons of Internalizing Problems in a Cohort of 8952 Adolescents from Five European Countries: The EU NET ADB Survey. Children, 10(1), 8. 10.3390/children10010008.

15. Ellins, J., Hocking, L., Al-Haboubi, M., Newbould, J., Fenton, S., Daniel, K., Stockwell, S., Leach, B., Sidhu, M., Bousfield, J., McKenna, G., Saunders, K., O’Neill, S., & Mays, N. (2023). Early Evaluation of the Children and Young People’s Mental Health Trailblazer programme: a rapid mixed-methods study. Available online at: file:///C:/Users/k1510035/Downloads/3041932%20(1).pdf. Last accessed: 5th May 2023.

16. Fergusson, D. M., Swain-Campbell, N. R., & Horwood, L. J. (2002). Does sexual violence contribute to elevated rates of anxiety and depression in females? [Article]. Psychological Medicine, 32(6), 991–996.

17. Ghavami, N., Katsiaficas, D., & Rogers, L. O. (2016). Toward an Intersectional Approach in Developmental Science: The Role of Race, Gender, Sexual Orientation, and Immigrant Status. Advances in Child Development and Behaviour, 50, 31–73.

18. Goodman, A., Lamping, D. L., & Ploubidis, G. B. (2010). When to use broader internalising and externalising subscales instead of the hypothesised five subscales on the Strengths and Difficulties Questionnaire (SDQ): data from British parents, teachers and children. Journal of Abnormal Child Psychology, 38(8), 1179–1191.

19. Goodman, A., Patel, V., & Leon, D. A. (2008). Child mental health differences amongst ethnic groups in Britain: a systematic review. BMC Public Health, 8(1), 258. 10.1186/1471-2458-8-258

20. Goodman, R. (2001). Psychometric properties of the strengths and difficulties questionnaire. Journal of the American Academy of Child and Adolescent Psychiatry, 40(11), 1337–1345.

21. Gutman, L. M. (2019). Do Conduct Problem Pathways Differ for Black and Minority Ethnic Children in the UK? An Examination of Trajectories from Early Childhood to Adolescence. Journal of Youth and Adolescence, 48(10), 1967–1979.

22. Harding, S., Read, U. M., Molaodi, O. R., Cassidy, A., Maynard, M. J., Lenguerrand, E., Astell-Burt, T., Teyhan, A., Whitrow, M., & Enayat, Z. E. (2015). The Determinants of young Adult Social well-being and Health (DASH) study: diversity, psychosocial determinants and health. Social Psychiatry and Psychiatric Epidemiology, 50(8), 1173–1188.

23. Hatch, S. L., Woodhead, C., Frissa, S., Fear, N. T., Verdecchia, M., Stewart, R., Reichenberg, A., Morgan, C., Bebbington, P., McManus, S., Brugha, T., Kankulu, B., Clark, J. L., Gazard, B., Medcalf, R., & Hotopf, M. (2012). Importance of thinking locally for mental health: data from cross-sectional surveys representing South East London and England. PLoS One, 7(12), e48012.

24. Hu, L. T., & Bentler, P. (1999). Cutoff criteria for fit indexes in covariance structure analysis: Conventional criteria versus new alternatives. Structural equation modeling: a multidisciplinary journal, 6(1), 1–55.

25. Jones, P. B. (2013). Adult mental health disorders and their age at onset. The British Journal of Psychiatry, 202(s54), s5–s10.

26. Karamanos, A., Mudway, I., Kelly, F., Beevers, S. D., Dajnak, D., Elia, C., Cruickshank, J. K., Lu, Y., Tandon, S., Enayat, E., Dazzan, P., Maynard, M., & Harding, S. (2021). Air pollution and trajectories of adolescent conduct problems: the roles of ethnicity and racism; evidence from the DASH longitudinal study. Social Psychiatry and Psychiatric Epidemiology, 56(11), 2029–2039.

27. Kern, M. R., Duinhof, E. L., Walsh, S. D., Cosma, A., Moreno-Maldonado, C., Molcho, M., Currie, C., & Stevens, G. (2020). Intersectionality and Adolescent Mental Well-being: A Cross-Nationally Comparative Analysis of the Interplay Between Immigration Background, Socioeconomic Status and Gender. Journal of Adolescent Health, 66(6s), S12–s20.

28. Kessler, R. C., Amminger, G. P., Aguilar-Gaxiola, S., Alonso, J., Lee, S., & Ustun, T. B. (2007). Age of onset of mental disorders: a review of recent literature. Current Opinion in Psychiatry, 20(4), 359–364.

29. Knowles, G., Gayer-Anderson, C., Beards, S., Blakey, R., Davis, S., Lowis, K., Stanyon, D., Ofori, A., Turner, A., Working Group, S., Pinfold, V., Bakolis, I., Reininghaus, U., Harding, S., & Morgan, C. (2021). Mental distress among young people in inner cities: the Resilience, Ethnicity and AdolesCent Mental Health (REACH) study. Journal of Epidemiology and Community Health, 75(6), 515–522.

30. Knowles, G., Gayer-Anderson, C., Blakey, R., Davis, S., Lowis, K., Stanyon, D., Ofori, A., Turner, A., Dorn, L., Beards, S., Pinfold, V., Reininghaus, U., Harding, S., & Morgan, C. (2022). Cohort Profile: Resilience, Ethnicity and AdolesCent mental Health (REACH). International Journal of Epidemiology, 51(5), e303–e313.

31. Knowles, G., Stanyon, D., Yamasaki, S., Miyashita, M., Gayer-Anderson, C., Endo, K., et al. (2023). Gender inequalities in trajectories of depressive symptoms among young people in London and Tokyo: a longitudinal cross-cohort study. PREPRINT. https://www.medrxiv.org/content/10.1101/2023.11.22.23298823v1.

32. Kuehner, C. (2017). Why is depression more common among women than among men? The Lancet Psychiatry, 4(2), 146–158.

33. Kuyken, W., Ball, S., Crane, C., Ganguli, P., Jones, B., Montero-Marin, J., Nuthall, E., Raja, A., Taylor, L., Tudor, K., Viner, R. M., Allwood, M., Aukland, L., Dunning, D., Casey, T., Dalrymple, N., De Wilde, K., Farley, E. R., Harper, J., Kappelmann, N., Kempnich, M., Lord, L., Medlicott, E., Palmer, L., Petit, A., Philips, A., Pryor-Nitsch, I., Radley, L., Sonley, A., Shackleford, J., Tickell, A., Blakemore, S. J., The Myriad Team, Ukoumunne, O. C., Greenberg, M. T., Ford, T., Dalgleish, T., Byford, S., & Williams, J. M. G. (2022). Effectiveness and cost-effectiveness of universal school-based mindfulness training compared with normal school provision in reducing risk of mental health problems and promoting well-being in adolescence: the MYRIAD cluster randomised controlled trial. Evidence Based Mental Health, 25(3), 99–109.

34. Lai, E., Wickham, S., Law, C., Whitehead, M., Barr, B., & Taylor-Robinson, D. (2019). Poverty dynamics and health in late childhood in the UK: evidence from the Millennium Cohort Study. Archives of Disease in Childhood, 104(11), 1049.

35. Maynard, M. J., Harding, S., & Minnis, H. (2007). Psychological well-being in Black Caribbean, Black African, and White adolescents in the UK Medical Research Council DASH study [journal article]. Social Psychiatry and Psychiatric Epidemiology, 42(9), 759–769.

36. McElroy, E., Tibber, M., Fearon, P., Patalay, P., & Ploubidis, G. B. (2023). Socioeconomic and sex inequalities in parent-reported adolescent mental ill-health: time trends in four British birth cohorts. Journal of Child Psychology and Psychiatry, 64(5), 758–767.

37. McManus, S., Bebbington, P., Jenkins, R., & Brugha, T. (2016). Mental health and wellbeing in England: Adult Psychiatric Morbidity Survey 2014. Available online at: https://digital.nhs.uk/data-and-information/publications/statistical/adult-psychiatric-morbidity-survey/adult-psychiatric-morbidity-survey-survey-of-mental-health-and-wellbeing-england-2014. Last accessed: 5th May 2023.

38. Montero-Marin, J., Allwood, M., Ball, S., Crane, C., De Wilde, K., Hinze, V., Jones, B., Lord, L., Nuthall, E., Raja, A., Taylor, L., Tudor, K., Blakemore, S. J., Byford, S., Dalgleish, T., Ford, T., Greenberg, M. T., Ukoumunne, O. C., Williams, J. M. G., & Kuyken, W. (2022). School-based mindfulness training in early adolescence: what works, for whom and how in the MYRIAD trial? Evidence Based Mental Health, 25(3), 117–124.

39. Morgan, C., Dazzan, P., Morgan, K., Jones, P., Harrison, G., Leff, J., Murray, R., & Fearon, P. (2006). First episode psychosis and ethnicity: initial findings from the AESOP study. World Psychiatry, 5(1), 40–46.

40. Morgan, C., Knowles, G., & Hutchinson, G. (2019). Migration, ethnicity and psychoses: evidence, models and future directions. World Psychiatry, 18(3), 247–258.

41. Murray, A. L., Ushakova, A., Speyer, L., Brown, R., Auyeung, B., & Zhu, X. (2022). Sex/gender differences in individual and joint trajectories of common mental health symptoms in early to middle adolescence. JCPP Advances, 2(1), e12057.

42. Muthén, L., & Muthén, B. (2017). Mplus User’s Guide. Eighth Edition.

43. Nazroo, J. Y., Bhui, K. S., & Rhodes, J. (2020). Where next for understanding race/ethnic inequalities in severe mental illness? Structural, interpersonal and institutional racism. Sociology of Health & Illness, 42(2), 262–276.

44. Office for National Statistics. (2012). Ethnicity and National Identity in England and Wales: 2011. Available online at: https://www.ons.gov.uk/peoplepopulationandcommunity/culturalidentity/ethnicity/articles/ethnicityandnationalidentityinenglandandwales/2012-12-11. Last accessed: 5th May 2023.

45. Patalay, P., & Demkowicz, O. (2023). Debate: Don’t mind the gap–why do we not care about the gender gap in common mental health difficulties? Child and Adolescent Mental Health.

46. Patalay, P., & Gage, S.H. (2019). Changes in millennial adolescent mental health and health-related behaviours over 10 years: a population cohort comparison study. International Journal of Epidemiology, 48(5), 1650–1664.

47. Patil, P. A., Porche, M. V., Shippen, N. A., Dallenbach, N. T., & Fortuna, L. R. (2018). Which girls, which boys? The intersectional risk for depression by race and ethnicity, and gender in the U.S. Clinical Psychology Review, 66, 51–68.

48. Patrick, R., Anstey, K., Lee, T., & Power, M. (2021). Fixing Lunch: The case for expanding free school meals. A Covid Realities and Child Poverty Action Group Rapid-Response Report. Available online at: https://cpag.org.uk/sites/default/files/files/policypost/Fixing_Lunch.pdf. Last accessed: 6th May 2023.

49. Piccinelli, M., & Wilkinson, G. (2000). Gender differences in depression: Critical review. The British Journal of Psychiatry, 177(6), 486–492.

50. Platt JM, Bates L, Jager J, McLaughlin KA, Keyes KM. Is the US Gender Gap in Depression Changing Over Time? A Meta-Regression. Am J Epidemiol. 2021 Jul 1;190(7):1190–1206. doi: 10.1093/aje/kwab002.

51. Platt, J., Prins, S., Bates, L., & Keyes, K. (2016). Unequal depression for equal work? How the wage gap explains gendered disparities in mood disorders. Social Science and Medicine, 149, 1–8.

52. Sadler, K., Vizard, T., Ford, T., Marcheselli, F., Pearce, N., Mandalia, D., Davis, J., Brodie, E., Forbes, N., Goodman, A., Goodman, R., McManus, S., & D., C. (2018). Mental Health of Children and Young People in England, 2017. Office for National Statistics: NHS Digital Retrieved from https://digital.nhs.uk/data-and-information/publications/statistical/mental-health-of-children-and-young-people-in-england/2017/2017. Last accessed: 4th May 2023.

53. Sellers, R., Warne, N., Pickles, A., Maughan, B., Thapar, A., & Collishaw, S. (2019). Cross-cohort change in adolescent outcomes for children with mental health problems. Journal of Child Psychology and Psychiatry, 60(7), 813–821.

54. Shuey, K. M., & Willson, A. E. (2008). Cumulative disadvantage and black-white disparities in life-course health trajectories. Research on aging, 30(2), 200–225.

55. Smith, N. R., Clark, C., Smuk, M., Cummins, S., & Stansfeld, S. A. (2015). The influence of social support on ethnic differences in well-being and depression in adolescents: findings from the prospective Olympic Regeneration in East London (ORiEL) study. Social Psychiatry and Psychiatric Epidemiology, 50(11), 1701–1711.

56. Stallard, P., Phillips, R., Montgomery, A. A., Spears, M., Anderson, R., Taylor, J., Araya, R., Lewis, G., Ukoumunne, O. C., Millings, A., Georgiou, L., Cook, E., & Sayal, K. (2013). A cluster randomised controlled trial to determine the clinical effectiveness and cost-effectiveness of classroom-based cognitive-behavioural therapy (CBT) in reducing symptoms of depression in high-risk adolescents. Health Technology Assessment, 17(47), vii–xvii, 1-109.

57. Office for National Statistics. (2022). Suicides in England and Wales: 2021 registrations. https://www.ons.gov.uk/peoplepopulationandcommunity/birthsdeathsandmarriages/deaths/bulletins/suicidesintheunitedkingdom/2021registrations#cite-this-statistical-bulletin. Last accessed: 5th May 2023.

58. Terhaag, S., Fitzsimons, E., Daraganova, G., & Patalay, P. (2021). Sex, ethnic and socioeconomic inequalities and trajectories in child and adolescent mental health in Australia and the UK: findings from national prospective longitudinal studies. Journal of Child Psychology and Psychiatry, 62(10), 1255–1267.

59. Wickham, S., Whitehead, M., Taylor-Robinson, D., & Barr, B. (2017). The effect of a transition into poverty on child and maternal mental health: a longitudinal analysis of the UK Millennium Cohort Study. Lancet Public Health, 2(3), e141–e148.

60. Xue, B., & McMunn, A. (2021). Gender differences in unpaid care work and psychological distress in the UK Covid-19 lockdown. PLoS One, 16(3), e0247959.

61. Yoon, Y., Eisenstadt, M., Lereya, S. T., & Deighton, J. (2022). Gender difference in the change of adolescents’ mental health and subjective wellbeing trajectories. European Child and Adolescent Psychiatry, 1–10.

62. Zilanawala, A., Sacker, A., & Kelly, Y. (2018). Mixed ethnicity and behavioural problems in the Millennium Cohort Study. Archives of Disease in Childhood, 103(1), 61–64.

