## Supplementary Information for "The emergence and persistence of inequalities in adolescent mental health: the Resilience, Ethnicity and AdolesCent Mental Health (REACH) cohorts"

**SUPPLEMENTARY MATERIAL**

### Figure S1. The accelerated (i.e., cohort-sequential) study design

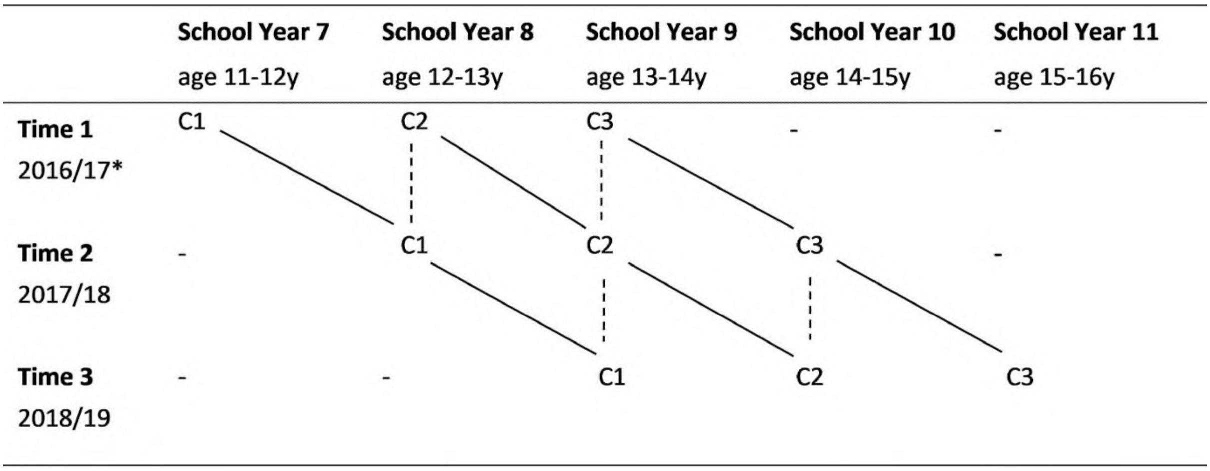

C1, Cohort 1; C2, Cohort 2; C3, Cohort 3. Dashed lines indicate points at which cohorts overlap. C1 and C2 overlap at School Year 8; C1, C2 and C3 overlap at School Year 9; and C2 and C3 overlap at School Year 10. *For one participating school, baseline data collection was deferred a year, so data were collected in 2018 (T1), 2019 (T2), 2020 (T3). Figure reproduced, with permission (Open Access) from Knowles et al., 2022.

**Appendix S1. Testing the assumptions of the accelerated study design.**

We performed a series of analyses to test the assumptions of the accelerated cohort study design (depicted in Figure S1), i.e., that the three cohorts are drawn from the same population and represent a common developmental trajectory from Year 7 to Year 11, and thus school year group, rather than wave of data collection, is the appropriate time metric for our analyses. For each distress metric, we (a) used t-tests and linear regression to compare mean distress scores at the overlapping segments (i.e., we compared Year 8 mean scores in Cohort 1 vs Cohort 2; Year 9 mean scores in Cohort 1 vs. Cohort 2 vs. Cohort 3; and Year 10 mean scores in Cohort 2 vs. Cohort 3); (b) fit a series of latent growth curve models (LCMs) within each cohort to check that the best-fitting functional form (i.e., shape of the trajectory) was consistent across cohorts (i.e., that a linear growth model fit similarly well in each cohort), and (c) fit multiple-group LCMs with cohort as the grouping variable and wave of data collection (i.e., wave 1, wave 2, wave 3) as the time metric, and constrained the factor means and variances to be equal across cohorts. Lack of evidence for a decline in model fit with the introduction of equality constraints – based on standard model fit indices, i.e., the CFI, TLI, RMSEA, SRMR, and $Chi^2$ LRT, collectively – suggests the three cohorts represent a common developmental trajectory from Year 7 to Year 11 and the overlapping segments of the cohorts can be combined to estimate trajectories over a 5-year period using 3 waves of data from each (Baer 2000, Duncan 2006). If supported, school year (Years 7-11) – rather than wave of data collection (waves 1-3) – would then be used as the time metric in all subsequent LCMs. As a final check, we also added 'cohort' as a covariate in unconditional LCMs, with school year as the time metric, to examine whether means and variances in growth parameters varied substantively by cohort (which would challenge the notion of a single developmental trend). Collectively, these analyses strongly supported the assumptions – i.e., there was little-to-no evidence of differences in year group specific mean distress scores by cohort, the linear growth model fit similarly well in each cohort, and there was no evidence of differences in growth parameters by cohort – so we proceeded to model a single developmental trajectory with school year as the time metric, i.e., 5 time points from Year 7 to Year 11 (as opposed to three separate trajectories, one per cohort, with wave of data collection as the time metric, i.e., 3 time points from T1 to T3).

**Figure S2. Distribution of distress scores, over time (i.e., by school year)**

**a) total difficulties** **b) internalising** **c) externalising**

**year 7**

**year 8**

**year 9**

**year 10**

**year 11**

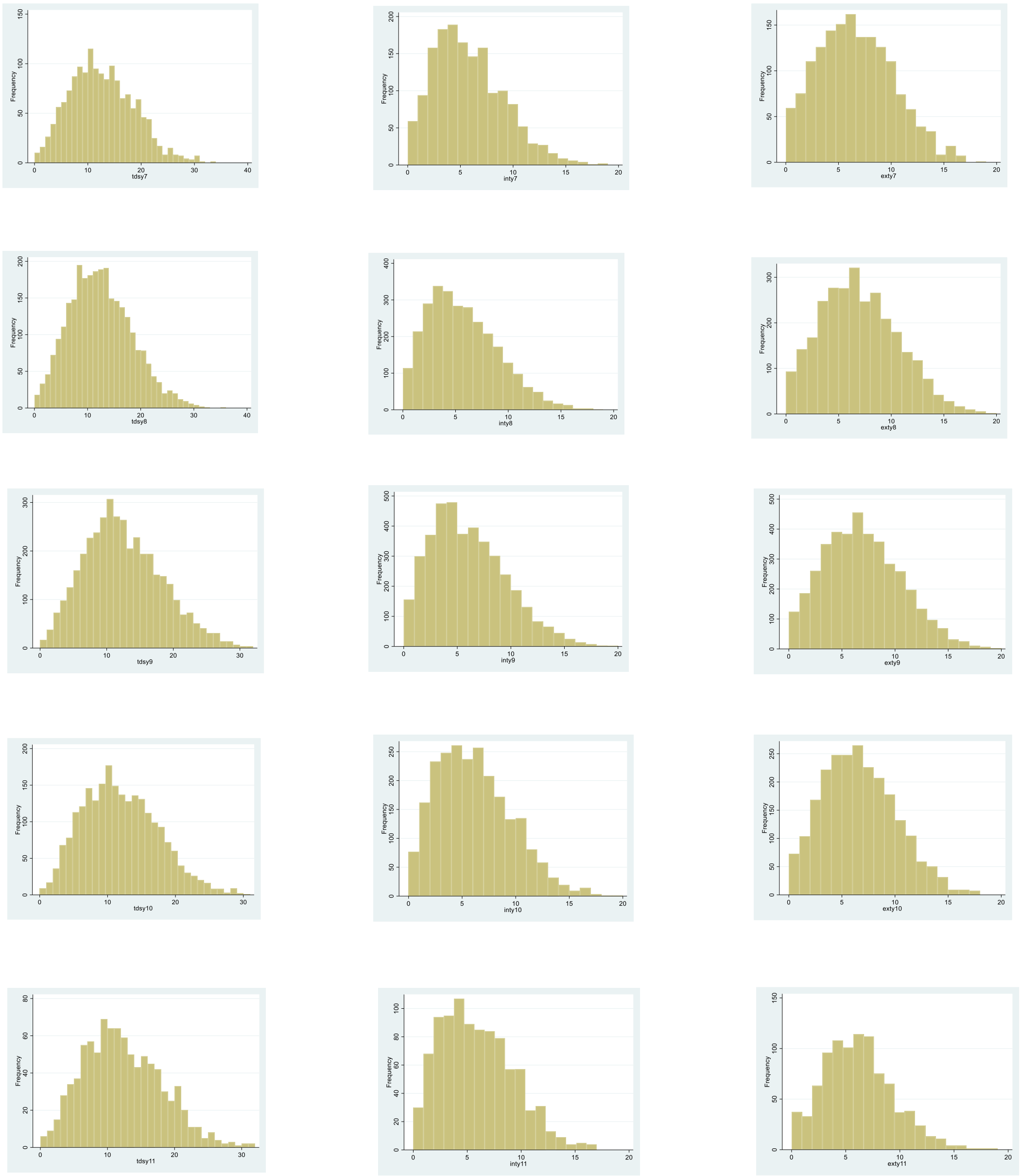

**Table S1. Model fit statistics.**

| Model | n | n free parameters | AIC | BIC | SSABIC | RMSEA (90% CI) | CFI | TLI | SRMR |
|---|---|---|---|---|---|---|---|---|---|
| **a) total difficulties** | | | | | | | | | |
| model 1 | 4652 | 10 | 69791.81 | 69856.26 | 69824.49 | 0.034 (0.025, 0.044) | 0.987 | 0.987 | 0.065 |
| model 2 | 4652 | 12 | 69622.77 | 69700.11 | 69661.98 | 0.030 (0.023, 0.039) | 0.988 | 0.986 | 0.060 |
| model 3 | 4588 | 12 | 69110.99 | 69188.16 | 69150.03 | 0.032 (0.025, 0.041) | 0.985 | 0.982 | 0.059 |
| model 4 | 4652 | 18 | 69737.28 | 69853.30 | 69796.10 | 0.021 (0.015, 0.027) | 0.988 | 0.984 | 0.048 |
| model 5 | 4588 | 22 | 68887.52 | 69029.00 | 68959.10 | 0.020 (0.015, 0.026) | 0.987 | 0.981 | 0.044 |
| **b) internalising** | | | | | | | | | |
| model 1 | 4662 | 10 | 58830.44 | 58894.91 | 58863.13 | 0.043 (0.034, 0.053) | 0.990 | 0.990 | 0.073 |
| model 2 | 4662 | 12 | 58394.74 | 58472.11 | 58433.98 | 0.036 (0.028, 0.044) | 0.994 | 0.992 | 0.066 |
| model 3 | 4597 | 12 | 58233.59 | 58310.78 | 58272.65 | 0.037 (0.029, 0.045) | 0.987 | 0.984 | 0.065 |
| model 4 | 4662 | 18 | 58768.09 | 58884.14 | 58826.95 | 0.027 (0.021, 0.033) | 0.983 | 0.976 | 0.053 |
| model 5 | 4597 | 22 | 57719.31 | 57860.84 | 57790.93 | 0.022 (0.017, 0.028) | 0.989 | 0.983 | 0.047 |
| **c) externalising** | | | | | | | | | |
| model 1 | 4660 | 10 | 59058.70 | 59123.16 | 59091.39 | 0.033 (0.024, 0.043) | 0.987 | 0.987 | 0.067 |
| model 2 | 4660 | 12 | 59043.90 | 59121.26 | 59083.13 | 0.030 (0.023, 0.039) | 0.985 | 0.982 | 0.061 |
| model 3 | 4595 | 12 | 58445.85 | 58523.04 | 58484.91 | 0.032 (0.025, 0.041) | 0.984 | 0.981 | 0.061 |
| model 4 | 4660 | 18 | 58978.34 | 59094.39 | 59037.19 | 0.021 (0.014, 0.027) | 0.987 | 0.982 | 0.050 |
| model 5 | 4595 | 22 | 58359.45 | 58500.97 | 58431.07 | 0.021 (0.016, 0.027) | 0.983 | 0.975 | 0.045 |

Model fit statistics for best-fitting latent curve models, i.e., linear growth LCMs with heteroscedastic residuals estimated using full information maximum likelihood, with robust standard errors to account for clustering at the school level. Fit statistics for other models not included in the final analysis (i.e., no-growth/intercept-only models; models with homoscedastic (rather than heteroscedastic) residuals; and models in which the time metric is wave of data collection rather than school year) are available on request. Model 1: unconditional; Model 2: model 1 + gender; Model 3: model 1 + free school meals status; Model 4: model 1 + ethnic group; Model 5: model 1 + gender + free school meals status + ethnic group. Abbreviations: AIC, Akaike's Information Criteria; BIC, Bayesian Information Criteria; SSABIC, Sample-Size Adjusted BIC; RMSEA, Root Mean Square Error of Approximation; CFI, Comparative Fit Index; TLI, Tucker Lewis Index; SRMR, Standardized Root Mean Squared Residual. Note: Satorra-Bentler likelihood ratio tests are highly sensitive to sample size and thus not recommended for large samples, so are not presented here but are available on request.

**Table S2. Growth parameter estimates (and 95% CIs) for interactions between (i) gender and FSM, (ii) ethnic group and FSM, and (iii) ethnic group and gender.**

| | Model A - gender * FSM | | Model B - ethnic group * FSM | | Model C - ethnic group * gender | | Model D – Ethnic group * FSM, within gender: Boys | | Model D – Ethnic group * FSM, within gender: Girls | |
|---|---|---|---|---|---|---|---|---|---|---|
| | b (95% CI) | p | b (95% CI) | p | b (95% CI) | p | b (95% CI) | p | b (95% CI) | p |
| **a) total difficulties** | | | | | | | | | | |
| **Factor means (reference group)** | | | | | | | | | | |
| **intercept** | 11.6 (11.1, 12.0) | <0.001 | 11.1 (10.2, 11.9) | <0.001 | 12.1 (11.5, 12.7) | <0.001 | 11.6 (10.8, 12.5) | <0.001 | 11.1 (10.2, 11.9) | <0.001 |
| **slope** | -0.15 (-0.28, -0.03) | 0.015 | -0.89 (-1.12, -0.65) | <0.001 | -0.27 (-0.48, -0.06) | 0.011 | -0.18 (-0.48, 0.12) | 0.250 | 0.49 (0.16, 0.81) | 0.003 |
| **Main effects** | | | | | | | | | | |
| **girl** | | | | | | | | | | |
| **intercept** | 0.29 (-0.40, 0.99) | 0.413 | 0.20 (-0.31, 0.71) | 0.432 | -0.82 (-1.96, 0.33) | 0.163 | - | - | - | - |
| **slope** | 0.64 (0.40, 0.88) | <0.001 | 0.70 (0.55, 0.86) | <0.001 | 0.88 (0.41, 1.36) | <0.001 | - | - | - | - |
| **fsm** | | | | | | | | | | |
| **intercept** | 0.85 (-0.02, 1.72) | 0.055 | 2.43 (0.80, 4.05) | 0.003 | 0.70 (0.11, 1.29) | 0.021 | 3.44 (1.48, 5.40) | 0.001 | 1.73 (-0.20, 3.65) | 0.078 |
| **slope** | -0.14 (-0.37, 0.10) | 0.248 | 0.01 (-0.61, 0.63) | 0.966 | -0.02 (-0.20, 0.17) | 0.860 | -0.64 (-1.42, 0.15) | 0.114 | 0.50 (-0.20, 1.21) | 0.163 |
| **black african** | | | | | | | | | | |
| **intercept** | -0.04 (-0.79, 0.70) | 0.909 | 0.34 (-0.51, 1.19) | 0.434 | -0.83 (-1.63, -0.02) | 0.044 | -0.32 (-1.39, 0.74) | 0.552 | 0.98 (-0.13, 2.08) | 0.083 |
| **slope** | -0.54 (-0.82, -0.27) | <0.001 | -0.52 (-0.78, -0.26) | <0.001 | -0.38 (-0.74, -0.01) | 0.043 | -0.43 (-0.83, -0.03) | 0.037 | -0.58 (-0.99, -0.17) | 0.006 |
| **black caribbean** | | | | | | | | | | |
| **intercept** | 0.99 (0.20, 1.78) | 0.014 | 1.56 (0.40, 2.73) | 0.009 | 0.06 (-0.81, 0.93) | 0.886 | 0.70 (-0.55, 1.94) | 0.273 | 2.42 (1.14, 3.69) | <0.001 |
| **slope** | -0.36 (-0.65, -0.07) | 0.016 | -0.36 (-0.71, -0.01) | 0.046 | -0.13 (-0.36, 0.09) | 0.245 | -0.26 (-0.75, 0.22) | 0.287 | -0.43 (-0.91, 0.04) | 0.074 |
| **mixed black and white** | | | | | | | | | | |
| **intercept** | 1.23 (0.39, 2.07) | 0.004 | 1.36 (0.68, 2.04) | <0.001 | 0.83 (-0.17, 1.84) | 0.105 | 0.74 (-0.74, 2.21) | 0.326 | 1.95 (0.44, 3.45) | 0.011 |
| **slope** | -0.46 (-0.79, -0.14) | 0.006 | -0.45 (-0.80, -0.10) | 0.012 | -0.60 (-1.05, -0.15) | 0.008 | -0.49 (-1.06, 0.08) | 0.093 | -0.37 (-0.95, 0.21) | 0.207 |
| **other ethnic group** | | | | | | | | | | |
| **intercept** | 0.28 (-0.37, 0.93) | 0.402 | 0.69 (-0.06, 1.44) | 0.073 | -0.10 (-1.21, 1.01) | 0.860 | 0.47 (-0.53, 1.47) | 0.357 | 0.93 (-0.11, 1.97) | 0.080 |
| **slope** | -0.14 (-0.34, 0.06) | 0.161 | -0.16 (-0.33, 0.00) | 0.055 | -0.07 (-0.34, 0.20) | 0.612 | -0.20 (-0.59, 0.18) | 0.293 | -0.11 (-0.50, 0.29) | 0.598 |
| **Interaction effects** | | | | | | | | | | |
| **girl*fsm** | | | | | | | | | | |
| **intercept** | -0.30 (-1.73, 1.14) | 0.688 | - | - | - | - | - | - | - | - |
| **slope** | 0.23 (-0.25, 0.72) | 0.346 | - | - | - | - | - | - | - | - |
| **ba*fsm** | | | | | | | | | | |
| **intercept** | - | - | -1.90 (-3.67, -0.13) | 0.036 | - | - | -2.91 (-5.25, -0.57) | 0.015 | -1.19 (-3.54, 1.15) | 0.317 |

| | | | | | | | | | | |
|---|---|---|---|---|---|---|---|---|---|---|
| **slope** | - | - | -0.11 (-0.68, 0.45) | 0.698 | - | - | 0.46 (-0.48, 1.39) | 0.338 | -0.52 (-1.38, 0.34) | 0.237 |
| **bc*fsm** | | | | | | | | | | |
| **intercept** | - | - | -2.37 (-4.37, -0.37) | 0.020 | - | - | -3.21 (-5.72, -0.70) | 0.012 | -1.88 (-4.36, 0.60) | 0.137 |
| **slope** | - | - | -0.04 (-0.69, 0.61) | 0.909 | - | - | 0.71 (-0.31, 1.73) | 0.173 | -0.58 (-1.51, 0.34) | 0.218 |
| **mxbw*fsm** | | | | | | | | | | |
| **intercept** | - | - | -1.18 (-2.55, 0.20) | 0.093 | - | - | -1.27 (-4.10, 1.56) | 0.379 | -1.34 (-4.15, 1.47) | 0.349 |
| **slope** | - | - | -0.07 (-0.50, 0.36) | 0.761 | - | - | 0.07 (-1.05, 1.18) | 0.904 | -0.07 (-1.13, 0.98) | 0.891 |
| **oth*fsm** | | | | | | | | | | |
| **intercept** | - | - | -2.02 (-3.76, -0.28) | 0.023 | - | - | -3.13 (-5.39, -0.87) | 0.007 | -1.24 (-3.48, 1.01) | 0.281 |
| **slope** | - | - | 0.05 (-0.61, 0.71) | 0.893 | - | - | 0.74 (-0.17, 1.65) | 0.109 | -0.49 (-1.31, 0.34) | 0.250 |
| **ba*girl** | | | | | | | | | | |
| **intercept** | - | - | - | - | 1.57 (0.35, 2.80) | 0.012 | - | - | - | - |
| **slope** | - | - | - | - | -0.33 (-0.87, 0.20) | 0.222 | - | - | - | - |
| **bc*girl** | | | | | | | | | | |
| **intercept** | - | - | - | - | 1.83 (-0.19, 3.85) | 0.076 | - | - | - | - |
| **slope** | - | - | - | - | -0.44 (-1.11, 0.23) | 0.201 | - | - | - | - |
| **mxbw*girl** | | | | | | | | | | |
| **intercept** | - | - | - | - | 0.81 (-0.88, 2.51) | 0.347 | - | - | - | - |
| **slope** | - | - | - | - | 0.27 (-0.57, 1.11) | 0.530 | - | - | - | - |
| **oth*girl** | | | | | | | | | | |
| **intercept** | - | - | - | - | 0.77 (-0.74, 2.29) | 0.317 | - | - | - | - |
| **slope** | - | - | - | - | -0.15 (-0.59, 0.29) | 0.501 | - | - | - | - |
| **b) internalising** | | | | | | | | | | |
| **Factor means (reference group)** | | | | | | | | | | |
| **intercept** | 5.25 (5.06, 5.44) | <0.001 | 4.40 (3.96, 4.84) | <0.001 | 5.28 (5.05, 5.52) | <0.001 | 5.10 (4.63, 5.57) | <0.001 | 5.90 (5.39, 6.41) | <0.001 |
| **slope** | -0.05 (-0.13, 0.04) | 0.270 | -0.54 (-0.67, -0.41) | <0.001 | -0.03 (-0.14, 0.07) | 0.519 | 0.03 (-0.15, 0.21) | 0.747 | 0.37 (0.17, 0.57) | <0.001 |
| **Main effects** | | | | | | | | | | |
| **girl** | | | | | | | | | | |
| **intercept** | 0.76 (0.43, 1.09) | <0.001 | 0.74 (0.52, 0.96) | <0.001 | 0.69 (0.30, 1.08) | 0.001 | - | - | - | - |
| **slope** | 0.51 (0.35, 0.67) | <0.001 | 0.50 (0.39, 0.61) | <0.001 | 0.47 (0.21, 0.74) | <0.001 | - | - | - | - |
| **fsm** | | | | | | | | | | |
| **intercept** | 0.38 (-0.11, 0.86) | 0.128 | 1.00 (-0.21, 2.22) | 0.106 | 0.34 (0.01, 0.67) | 0.045 | 1.44 (0.30, 2.58) | 0.013 | 0.67 (-0.49, 1.83) | 0.256 |
| **slope** | -0.03 (-0.19, 0.13) | 0.725 | -0.06 (-0.52, 0.40) | 0.793 | -0.04 (-0.14, 0.06) | 0.440 | -0.45 (-0.92, 0.02) | 0.060 | 0.25 (-0.18, 0.69) | 0.254 |
| **black african** | | | | | | | | | | |
| **intercept** | -0.47 (-0.84, -0.09) | 0.015 | -0.35 (-0.81, 0.11) | 0.134 | -0.58 (-0.99, -0.17) | 0.006 | -0.37 (-0.99, 0.25) | 0.247 | -0.34 (-1.00, 0.33) | 0.317 |
| **slope** | -0.32 (-0.47, -0.17) | <0.001 | -0.31 (-0.45, -0.18) | <0.001 | -0.32 (-0.49, -0.15) | <0.001 | -0.41 (-0.65, -0.17) | 0.001 | -0.20 (-0.46, 0.05) | 0.118 |

| | | | | | | | | | | |
|---|---|---|---|---|---|---|---|---|---|---|
| **black caribbean** | | | | | | | | | | |
| **intercept** | -0.53 (-0.93, -0.13) | 0.010 | -0.20 (-0.87, 0.47) | 0.556 | -0.76 (-1.10, -0.41) | <0.001 | -0.41 (-1.14, 0.32) | 0.266 | -0.03 (-0.79, 0.74) | 0.950 |
| **slope** | -0.08 (-0.28, 0.13) | 0.448 | -0.12 (-0.36, 0.11) | 0.306 | -0.03 (-0.19, 0.13) | 0.713 | -0.14 (-0.43, 0.15) | 0.354 | -0.08 (-0.37, 0.21) | 0.601 |
| **mixed black and white** | | | | | | | | | | |
| **intercept** | -0.15 (-0.53, 0.23) | 0.427 | -0.06 (-0.50, 0.38) | 0.776 | -0.09 (-0.43, 0.24) | 0.589 | -0.22 (-1.08, 0.64) | 0.620 | 0.07 (-0.84, 0.98) | 0.880 |
| **slope** | -0.22 (-0.39, -0.05) | 0.010 | -0.23 (-0.41, -0.05) | 0.014 | -0.35 (-0.56, -0.14) | 0.001 | -0.30 (-0.64, 0.04) | 0.082 | -0.13 (-0.48, 0.23) | 0.490 |
| **other ethnic group** | | | | | | | | | | |
| **intercept** | 0.02 (-0.35, 0.39) | 0.911 | 0.14 (-0.29, 0.57) | 0.522 | 0.12 (-0.45, 0.69) | 0.676 | 0.33 (-0.25, 0.92) | 0.262 | -0.04 (-0.66, 0.59) | 0.911 |
| **slope** | -0.15 (-0.32, 0.03) | 0.102 | -0.14 (-0.28, -0.01) | 0.041 | -0.17 (-0.35, 0.02) | 0.076 | -0.26 (-0.49, -0.04) | 0.023 | -0.01 (-0.25, 0.23) | 0.929 |
| **Interaction effects** | | | | | | | | | | |
| **girl*fsm** | | | | | | | | | | |
| **intercept** | -0.07 (-0.81, 0.66) | 0.846 | - | - | - | - | - | - | - | - |
| **slope** | -0.02 (-0.27, 0.22) | 0.865 | - | - | - | - | - | - | - | - |
| **ba*fsm** | | | | | | | | | | |
| **intercept** | - | - | -0.63 (-2.01, 0.75) | 0.373 | - | - | -1.22 (-2.58, 0.14) | 0.079 | -0.16 (-1.57, 1.25) | 0.822 |
| **slope** | - | - | -0.01 (-0.50, 0.47) | 0.961 | - | - | 0.51 (-0.05, 1.07) | 0.074 | -0.44 (-0.97, 0.09) | 0.105 |
| **bc*fsm** | | | | | | | | | | |
| **intercept** | - | - | -1.22 (-2.77, 0.34) | 0.125 | - | - | -1.59 (-3.05, -0.12) | 0.034 | -0.95 (-2.44, 0.54) | 0.212 |
| **slope** | - | - | 0.13 (-0.45, 0.72) | 0.657 | - | - | 0.55 (-0.06, 1.15) | 0.077 | -0.20 (-0.77, 0.37) | 0.486 |
| **mxbw*fsm** | | | | | | | | | | |
| **intercept** | - | - | -0.57 (-1.63, 0.48) | 0.285 | - | - | -0.28 (-1.93, 1.38) | 0.743 | -0.95 (-2.64, 0.74) | 0.271 |
| **slope** | - | - | 0.04 (-0.31, 0.38) | 0.842 | - | - | 0.12 (-0.55, 0.79) | 0.724 | 0.01 (-0.64, 0.66) | 0.972 |
| **oth*fsm** | | | | | | | | | | |
| **intercept** | - | - | -0.65 (-1.83, 0.53) | 0.281 | - | - | -1.25 (-2.57, 0.06) | 0.062 | -0.20 (-1.56, 1.15) | 0.768 |
| **slope** | - | - | -0.01 (-0.50, 0.49) | 0.977 | - | - | 0.55 (0.01, 1.09) | 0.047 | -0.47 (-0.97, 0.05) | 0.074 |
| **ba*girl** | | | | | | | | | | |
| **intercept** | - | - | - | - | 0.22 (-0.16, 0.61) | 0.252 | - | - | - | - |
| **slope** | - | - | - | - | 0.01 (-0.29, 0.30) | 0.972 | - | - | - | - |
| **bc*girl** | | | | | | | | | | |
| **intercept** | - | - | - | - | 0.44 (-0.57, 1.46) | 0.393 | - | - | - | - |
| **slope** | - | - | - | - | -0.09 (-0.48, 0.31) | 0.668 | - | - | - | - |
| **mxbw*girl** | | | | | | | | | | |
| **intercept** | - | - | - | - | -0.12 (-0.80, 0.56) | 0.721 | - | - | - | - |
| **slope** | - | - | - | - | 0.26 (-0.20, 0.72) | 0.264 | - | - | - | - |
| **oth*girl** | | | | | | | | | | |
| **intercept** | - | - | - | - | -0.20 (-0.84, 0.45) | 0.549 | - | - | - | - |

| | | | | | | | | | | |
|---|---|---|---|---|---|---|---|---|---|---|
| slope | - | - | - | - | 0.04 (-0.22, 0.30) | 0.752 | - | - | - | - |
| **c) externalising** | | | | | | | | | | |
| **Factor means (reference group)** | | | | | | | | | | |
| intercept | 6.30 (5.90, 6.69) | <0.001 | 6.68 (6.05, 7.31) | <0.001 | 6.76 (6.37, 7.15) | <0.001 | 6.51 (6.00, 7.02) | <0.001 | 5.2 (4.7, 5.7) | <0.001 |
| slope | -0.09 (-0.18, -0.01) | 0.031 | -0.32 (-0.48, -0.17) | <0.001 | -0.21 (-0.33, -0.09) | <0.001 | -0.19 (-0.37, -0.01) | 0.042 | 0.10 (-0.09, 0.30) | 0.303 |
| **Main effects** | | | | | | | | | | |
| girl | | | | | | | | | | |
| intercept | -0.47 (-0.94, 0.01) | 0.054 | -0.54 (-0.88, -0.21) | 0.002 | -1.41 (-2.28, -0.55) | 0.001 | - | - | - | - |
| slope | 0.13 (0.01, 0.24) | 0.028 | 0.19 (0.10, 0.27) | <0.001 | 0.38 (0.15, 0.60) | 0.001 | - | - | - | - |
| fsm | | | | | | | | | | |
| intercept | 0.51 (-0.07, 1.09) | 0.083 | 1.37 (0.74, 2.01) | <0.001 | 0.37 (0.01, 0.74) | 0.047 | 1.80 (0.56, 3.04) | 0.004 | 1.08 (-0.10, 2.27) | 0.074 |
| slope | -0.10 (-0.30, 0.09) | 0.289 | 0.10 (-0.15, 0.36) | 0.427 | 0.02 (-0.13, 0.16) | 0.837 | -0.10 (-0.58, 0.38) | 0.687 | 0.25 (-0.18, 0.67) | 0.254 |
| black african | | | | | | | | | | |
| intercept | 0.48 (0.04, 0.93) | 0.034 | 0.74 (0.27, 1.22) | 0.002 | -0.11 (-0.61, 0.39) | 0.654 | 0.14 (-0.53, 0.82) | 0.676 | 1.34 (0.66, 2.02) | <0.001 |
| slope | -0.24 (-0.37, -0.10) | 0.001 | -0.21 (-0.35, -0.08) | 0.002 | -0.09 (-0.32, 0.14) | 0.459 | -0.04 (-0.28, 0.21) | 0.779 | -0.38 (-0.63, -0.13) | 0.003 |
| black caribbean | | | | | | | | | | |
| intercept | 1.49 (0.92, 2.06) | <0.001 | 1.77 (1.09, 2.46) | <0.001 | 0.87 (0.27, 1.46) | 0.004 | 1.13 (0.34, 1.92) | 0.005 | 2.41 (1.62, 3.19) | <0.001 |
| slope | -0.27 (-0.43, -0.11) | 0.001 | -0.23 (-0.43, -0.03) | 0.022 | -0.10 (-0.30, 0.10) | 0.326 | -0.12 (-0.41, 0.18) | 0.439 | -0.34 (-0.63, -0.06) | 0.019 |
| mixed black and white | | | | | | | | | | |
| intercept | 1.45 (0.78, 2.12) | <0.001 | 1.49 (0.75, 2.23) | <0.001 | 1.07 (0.36, 1.79) | 0.003 | 1.14 (0.21, 2.08) | 0.017 | 1.85 (0.92, 2.78) | <0.001 |
| slope | -0.27 (-0.47, -0.07) | 0.010 | -0.25 (-0.48, -0.01) | 0.038 | -0.30 (-0.55, -0.05) | 0.018 | -0.27 (-0.62, 0.08) | 0.127 | -0.22 (-0.57, 0.13) | 0.211 |
| other ethnic group | | | | | | | | | | |
| intercept | 0.27 (-0.18, 0.72) | 0.241 | 0.52 (0.08, 0.96) | 0.022 | -0.17 (-0.85, 0.52) | 0.631 | 0.11 (-0.53, 0.75) | 0.733 | 0.93 (0.29, 1.57) | 0.004 |
| slope | 0.01 (-0.10, 0.11) | 0.890 | 0.00 (-0.14, 0.14) | 0.999 | 0.09 (-0.09, 0.27) | 0.315 | 0.08 (-0.15, 0.31) | 0.501 | -0.07 (-0.31, 0.16) | 0.538 |
| **Interaction effects** | | | | | | | | | | |
| girl*fsm | | | | | | | | | | |
| intercept | -0.27 (-1.23, 0.68) | 0.577 | - | - | - | - | - | - | - | - |
| slope | 0.23 (-0.11, 0.57) | 0.187 | - | - | - | - | - | - | - | - |
| ba*fsm | | | | | | | | | | |
| intercept | | - | -1.22 (-1.79, -0.64) | <0.001 | - | - | -1.44 (-2.92, 0.04) | 0.056 | -1.12 (-2.56, 0.32) | 0.126 |
| slope | | - | -0.14 (-0.30, 0.02) | 0.089 | - | - | -0.16 (-0.73, 0.41) | 0.573 | -0.07 (-0.59, 0.44) | 0.782 |
| bc*fsm | | | | | | | | | | |
| intercept | | - | -1.22 (-2.01, -0.43) | 0.003 | - | - | -1.46 (-3.05, 0.13) | 0.071 | -1.13 (-2.65, 0.40) | 0.149 |
| slope | | - | -0.16 (-0.43, 0.11) | 0.244 | - | - | 0.10 (-0.53, 0.72) | 0.764 | -0.34 (-0.90, 0.22) | 0.230 |
| mxbw*fsm | | | | | | | | | | |
| intercept | | - | -0.56 (-1.22, 0.10) | 0.096 | - | - | -0.90 (-2.70, 0.89) | 0.323 | -0.37 (-2.10, 1.36) | 0.678 |

| | | | | | | | | | |
|---|---|---|---|---|---|---|---|---|---|
| **slope** | - | -0.12 (-0.43, 0.18) | 0.426 | - | - | -0.05 (-0.73, 0.63) | 0.889 | -0.14 (-0.78, 0.49) | 0.660 |
| **oth*fsm** | | | | | | | | | |
| **intercept** | - | -1.20 (-1.90, -0.50) | 0.001 | - | - | -1.50 (-2.93, -0.07) | 0.040 | -1.01 (-2.39, 0.38) | 0.153 |
| **slope** | - | -0.02 (-0.29, 0.25) | 0.865 | - | - | 0.06 (-0.49, 0.61) | 0.827 | -0.07 (-0.56, 0.43) | 0.798 |
| **ba*girl** | | | | | | | | | |
| **intercept** | - | - | - | 1.20 (0.24, 2.16) | 0.014 | - | - | - | - |
| **slope** | - | - | - | -0.30 (-0.61, 0.00) | 0.048 | - | - | - | - |
| **bc*girl** | | | | | | | | | |
| **intercept** | - | - | - | 1.25 (0.12, 2.38) | 0.031 | - | - | - | - |
| **slope** | - | - | - | -0.33 (-0.64, -0.02) | 0.035 | - | - | - | - |
| **mxbw*girl** | | | | | | | | | |
| **intercept** | - | - | - | 0.77 (-0.42, 1.95) | 0.203 | - | - | - | - |
| **slope** | - | - | - | 0.06 (-0.34, 0.45) | 0.768 | - | - | - | - |
| **oth*girl** | | | | | | | | | |
| **intercept** | - | - | - | 0.89 (-0.26, 2.03) | 0.129 | - | - | - | - |
| **slope** | - | - | - | -0.18 (-0.43, 0.07) | 0.155 | - | - | - | - |

Abbreviations: CI, confidence intervals; b, unstandardised coefficients; BA, Black African; BC, Black Caribbean; MXBW, mixed black-and-white; OTH, other ethnic group; FSM, free school meals. To aid interpretation, please see Figure 3 for visual representation of the estimated mean trajectories for each intersecting-identity group, created using 'loop plots' function in Mplus.

**Table S3. Sensitivity analysis: estimates (and 95% CIs) from linear latent growth curve models with school fitted as fixed effect.**

| | Model 2 - gender | Model 3 - FSM | Model 4 - ethnic group | Model 5 - gender + FSM + ethnic group |
|---|---|---|---|---|
| | b (95% CI) | b (95% CI) | b (95% CI) | b (95% CI) |
| **a) total difficulties** | | | | |
| **girl** | | | | |
| **intercept** | 0.22 (-0.27, 0.71) | - | - | 0.20 (-0.29, 0.69) |
| **slope** | 0.70 (0.51, 0.89) | - | - | 0.69 (0.50, 0.88) |
| **receiving free school meals** | | | | |
| **intercept** | - | 0.66 (0.18, 1.14) | - | 0.59 (0.11, 1.07) |
| **slope** | - | 0.05 (-0.13, 0.24) | - | 0.06 (-0.13, 0.24) |
| **black african** | | | | |
| **intercept** | - | - | 0.01 (-0.73, 0.75) | -0.07 (-0.81, 0.67) |
| **slope** | - | - | -0.53 (-0.81, -0.25) | -0.52 (-0.80, -0.24) |
| **black caribbean** | | | | |
| **intercept** | - | - | 1.15 (0.35, 1.95) | 1.03 (0.22, 1.83) |
| **slope** | - | - | -0.35 (-0.66, -0.04) | -0.35 (-0.66, -0.04) |
| **mixed black and white** | | | | |
| **intercept** | - | - | 1.37 (0.47, 2.27) | 1.27 (0.37, 2.17) |
| **slope** | - | - | -0.44 (-0.78, -0.10) | -0.46 (-0.81, -0.12) |
| **other ethnic group** | | | | |
| **intercept** | - | - | 0.35 (-0.32, 1.02) | 0.30 (-0.37, 0.97) |
| **slope** | - | - | -0.14 (-0.39, 0.12) | -0.14 (-0.39, 0.12) |
| **b) internalising** | | | | |
| **girl** | | | | |
| **intercept** | 0.82 (0.53, 1.11) | - | - | 0.81 (0.52, 1.10) |
| **slope** | 0.50 (0.39, 0.62) | - | - | 0.50 (0.39, 0.62) |
| **receiving free school meals** | | | | |
| **intercept** | | 0.20 (-0.09, 0.48) | - | 0.23 (-0.06, 0.51) |
| **slope** | | 0.03 (-0.09, 0.14) | - | 0.01 (-0.10, 0.13) |
| **black african** | | | | |
| **intercept** | - | - | -0.56 (-1.00, -0.12) | -0.53 (-0.97, -0.09) |

| | | | | |
|---|---|---|---|---|
| **slope** | - | - | -0.29 (-0.46, -0.12) | -0.29 (-0.46, -0.12) |
| **black caribbean** | | | | |
| **intercept** | - | - | -0.61 (-1.09, -0.13) | -0.61 (-1.09, -0.13) |
| **slope** | - | - | -0.03 (-0.22, 0.16) | -0.05 (-0.23, 0.14) |
| **mixed black and white** | | | | |
| **intercept** | - | - | -0.17 (-0.70, 0.37) | -0.20 (-0.73, 0.34) |
| **slope** | - | - | -0.19 (-0.40, 0.03) | -0.20 (-0.41, 0.01) |
| **other ethnic group** | | | | |
| **intercept** | - | - | -0.04 (-0.43, 0.36) | -0.02 (-0.42, 0.38) |
| **slope** | - | - | -0.11 (-0.27, 0.04) | -0.13 (-0.28, 0.03) |
| **c) externalising** | | | | |
| **girl** | | | | |
| **intercept** | -0.60 (-0.91, -0.29) | - | - | -0.60 (-0.91, -0.30) |
| **slope** | 0.17 (0.06, 0.29) | - | - | 0.16 (0.05, 0.28) |
| **receiving free school meals** | | | | |
| **intercept** | - | 0.49 (0.19, 0.79) | - | 0.38 (0.08, 0.68) |
| **slope** | - | 0.01 (-0.10, 0.12) | - | 0.03 (-0.08, 0.14) |
| **black african** | | | | |
| **intercept** | - | - | 0.63 (0.17, 1.09) | 0.53 (0.07, 0.99) |
| **slope** | - | - | -0.26 (-0.43, -0.09) | -0.25 (-0.42, -0.08) |
| **black caribbean** | | | | |
| **intercept** | - | - | 1.74 (1.24, 2.25) | 1.63 (1.12, 2.13) |
| **slope** | - | - | -0.32 (-0.50, -0.13) | -0.30 (-0.49, -0.11) |
| **mixed black and white** | | | | |
| **intercept** | - | - | 1.61 (1.05, 2.18) | 1.55 (0.98, 2.11) |
| **slope** | - | - | -0.29 (-0.50, -0.08) | -0.29 (-0.50, -0.09) |
| **other ethnic groups** | | | | |
| **intercept** | - | - | 0.41 (-0.01, 0.83) | 0.35 (-0.07, 0.77) |
| **slope** | - | - | -0.03 (-0.18, 0.13) | -0.02 (-0.17, 0.14) |

School fitted as a fixed effect (i.e., covariate/independent variable). FSM, free school meals. Estimates are unstandardised.

**Figure S3. Estimated mean distress trajectories (with 95% CIs) at the intersections of ethnic group and free school meals status, within gender.**

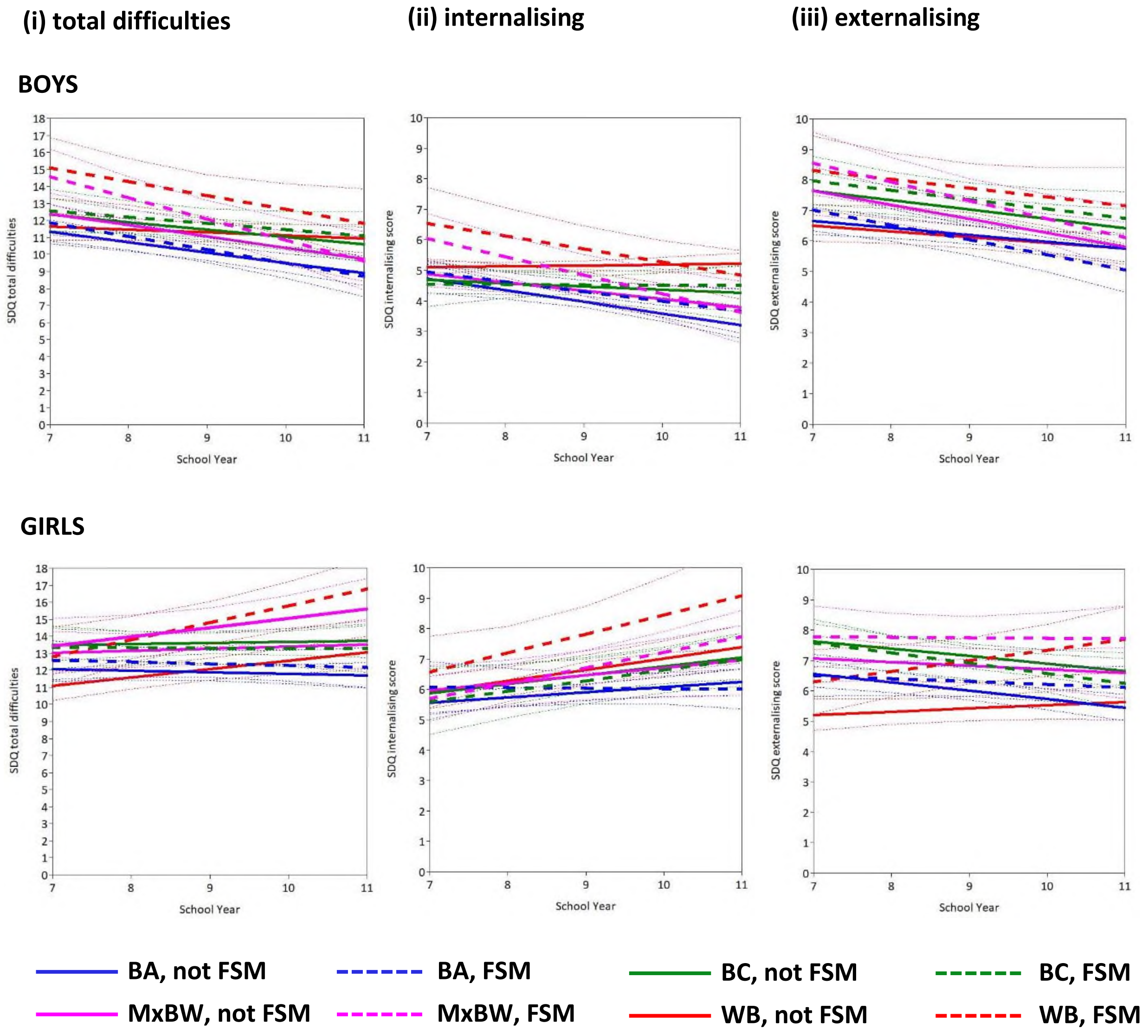

Bold lines: estimated mean trajectory per intersecting-identity group. Faint dashed lines: 95% confidence intervals. Abbreviations: FSM, free school meals; BA, Black African; BC, Black Caribbean; MxBW, Mixed Black-and-White; WB, White British. Note: 'other' ethnic group omitted to aid interpretation of figures, but parameter estimates for all groups – and for interaction terms – are provided in Table S2.

**Appendix S2. Young persons' perspectives.**

This work is coproduced with five young people (co-authors TR, AH, KSCG, JK and NT) from inner-London schools who work on REACH as (paid, part-time) co-researchers. Their reflections on this work are provided in their own words, below. For context, all five young co-researchers are 18-19 years old at the time of writing and have been working on reach for the last 2-4 years.

**Our reflections**

**(1) on gender**

There is a difference in internalising scores between boys and girls, and this difference gets bigger with age. We were not surprised to see this because girls tend to have more social pressures than boys. But we didn't expect the gap to be quite so wide and we were surprised to see the slope decreasing, rather than increasing a bit, for boys, because some of the same pressures that girls face also affect boys (e.g., pressures related to exams and grades). We speculate that this may be because there is still a lot of stigma around boys admitting to difficulties with mental health, like 'you should be a man', so boys are more likely to hide it. Boys may feel pressure to conform to traditional masculine gender roles, which can discourage them from seeking mental health support and make them more likely to internalise their emotions. This can make boys less likely to admit to mental health distress and more likely to suffer in silence. Also, we think that boys may be more likely to face criticism and judgment for expressing emotions or seeking help. It'd be interesting to know what causes this gender difference and whether asking boys about their 'stress levels' (rather than sad, low, anxious, etc) is more effective in getting them to open up.

We noticed that there was a big difference in the way boys and girls dealt with their feelings. Girls had a more challenging time, especially as we progressed through education. As girls, we tend to be aware of and worry about the consequences of underperforming in school and maybe even disappointing people based on their perception of our performance. We were surprised to see that the gap between boys and girls got smaller for some things, even though we thought boys would have more of the same problems that girls do.

For behaviour, some of us think it makes sense to see this pattern between boys and girls, because boys start to take school more seriously and stop 'playing around' as they get older. For girls, it'd be interesting, if we had more data points, to see if it gets worse and then improves again, because behaviour is usually good in year 7, gets worse in years 8 and 9 as you get more confident, and then improves again for GCSEs when things get serious.

**(2) on money, free school meals**

We found it surprising that the difference between those who get FSM and those who don’t wasn’t larger. We thought there’d be a bigger gap. Being from a low-income family can have a significant impact on young people because they might not be able to afford holidays, uniform, go on school trips, or have access to vital space or technology to complete tasks like homework and buy new things, which could result in social marginalisation and make you stand out. When you're older, the difference will be less noticeable because you'll be able to find employment and earn money on your own without needing to rely on your family. Although, coming from a low-income family can have an impact on your future employment because some positions require specific experiences that you were unable to obtain because of your household income.

Maybe the gap is small because students aren't open about their financial struggles or because other factors, such as social media, friendships and exams may have a greater impact on mental health at our age. Or maybe it’s because a lot of the young people who are barely above the FSM margin but are still not considered FSM, are still really struggling for money and are still having difficulties, which could possibly make the difference look smaller. In our schools, you don’t really feel there’s that much difference in terms of how much money everyone has – you only see that on the way to and from school, when you’re going through rich areas etc, whereas most of our friends at school are similar to us. However, there are instances in school where the distinctions are made clear, usually during school trips when our peers are called to the front to get FSM pack lunches, which may cause them to feel alienated.

**(3) on ethnic group, race – and behaviour**

We think it’s really important to consider how behaviour is measured and defined, who in society (and in schools) makes the rules, and what that means for young people from different races and ethnicities and for people who don’t have much money. Ethnicity plays a large role in young people’s experiences within schools, which can also have a significant impact and bearing on our mental health.

The issue of over-policing of mixed-race and black students in schools is significant. We see it a lot in schools’ behaviour policies – rules that aren’t technically “meant” to be targeted at anyone, but actually do end up discriminating against Black kids. For example, we’re told in the rules not to get “extreme” haircuts or hairstyles – but what counts as extreme? Like if you look at the behaviour policies, they don’t explicitly refer to us, but they include stuff like hair styles that are way more common in Black young people. Most times, it’s traditional Black hairstyles like braids that are seen as such. And these unfair rules lead to punishments such as detention and internal exclusion. It’s the failure of the institution, not the young people; we are effectively being punished for our culture.

As Black people, we are labelled for breaking the rules, but the rules are wrong. In mental health research, this is a problem because when completing questionnaires ourselves, and doing it honestly, we have to tick 'yes' to things like 'do you get in trouble a lot' because the rules are designed to get us in trouble. This means we get told off more, which then in turn means our answer to the question is 'yes' even though there's nothing wrong with our behaviour. We think this could explain some of the patterns in the results, like the 'worse behaviour' in Black Caribbean and Mixed Black-and-White kids in the first few years of secondary school. Although, for Mixed Black-and-White kids, it could be related to 'identity crisis' – like, you're 'too white to be black, too black to be white' – which can be difficult, especially when you're younger and you're trying to fit in. Also, as a young Black person, you become more and more aware of the injustices in society when you start secondary school – you see it and feel it more – and it gets to you. Maybe this is why Black Caribbean and Mixed-Black and-White kids are doing worse at the start in terms of 'behaviour'.

The data also shows strong evidence of higher initial levels of general distress among Black Caribbean and Mixed Black and White groups compared with Black African and White British groups. We believe there are many reasons for this, the most important being cultural reasons. In many Black African and Black Caribbean households, discussions around mental health are generally not encouraged, meaning young people from these backgrounds may be forced to internalise their emotions, which could lead to distress that sometimes results in behaviour issues at school for example. This could be a reason as to why there are some differences in the internalising patterns versus the externalising patterns.

On the other hand, the patterns for Black African young people (i.e., that they seem to be doing better, overall) and how much they differ from not just the white British group but also the Black Caribbean group are really interesting. It also might be due to cultural factors. For example, there might be things in their culture that help them do well, e.g., close families and communities, 'getting perspective' from older generations ('if you have a roof over your head, you're fine'), etc. We need to do more research to understand why we see this trend for Black African, Black Caribbean, and Mixed-black-and-white young people: we need to know if we can take it at face value or if there are other reasons we're seeing these patterns – because, if it's due to other reasons, we're at risk of underestimating problems and assuming particular groups are fine when maybe they're not. We need more research on this.
